# EXHEART: A Fairness-Aware Explainable Stacked Ensemble for Cardiovascular Disease Classification with Cross-Instrument Disparity Attribution

**DOI:** 10.64898/2026.06.03.26354879

**Authors:** Md Anas Biswas, Alif Laila

**Affiliations:** School of Computing, University of Portsmouth, Portsmouth, UK

**Keywords:** Heart disease classification, Machine learning, Explainability, SHAP, LIME, Fairness, Calibration, BRFSS, Stacked ensemble, Temporal drift

## Abstract

**Background:** Machine learning models trained on population health surveys offer scalable tools for cardiovascular screening, but recurring methodological weaknesses undermine their credibility and equity: data leakage from synthetic oversampling, qualitative rather than quantitative explainability evaluation, and the absence of demographic fairness auditing at the clinical operating threshold.

**Methods:** We present EXHEART, a leakage-free stacked ensemble pipeline trained on BRFSS 2015 (n = 253,680) and validated on BRFSS 2020 (n = 319,795; temporal transport and retrain) and a clinical cardiovascular examination dataset (n = 68,730). The pipeline combines XGBoost, LightGBM, Random Forest, and a multi-layer perceptron as base learners with 5-fold out-of-fold logistic regression stacking and Platt scaling calibration. A quantitative SHAP–LIME consistency framework, based on Kendall-τ rank correlation and Jaccard overlap, accompanies a decision-curve analysis, a subgroup-stratified SHAP interaction analysis, and an intersectional fairness audit (Sex × Age × Income) with threshold-shifting mitigation and a frontier of the fairness–utility trade-off. The framework also adds cross-instrument fairness-disparity attribution, an empirical diagnostic that provides evidence on whether an observed subgroup disparity is more consistent with a measurement-induced or a substantive explanation by re-validating it on a dataset that measures the same clinical construct objectively. On heart disease, this diagnostic associates 89% of the sex TPR gap (95% CI [0.65, 0.99]) with the self-reported survey outcome rather than with a substantive risk difference.

**Results:** On BRFSS 2015, EXHEART achieves AUC-ROC = 0.850, AUPRC = 0.371, Brier score = 0.071, and reduces ECE by 96% (0.256 → 0.011) via Platt scaling. Global SHAP–LIME rank agreement is moderate-to-strong (Kendall-τ = 0.580, Spearman-ρ = 0.818) with a substantial top-3 divergence (Jaccard@3 = 0.200), where Stroke flips from SHAP rank 8 to LIME rank 1. The Sex TPR gap is 0.124 at the screening threshold; intersectional Sex × Age disparities reach 0.649 among adequately-powered cells, 5.2× the single-attribute gap. Temporal transport to BRFSS 2020 collapses sensitivity from 0.776 to 0.267, while retraining restores AUC = 0.840 and ECE = 0.012. On clinical examination data, the Sex TPR gap collapses to 0.014; the attribution test indicates this gap is instrument-dependent, consistent with a measurement or outcome-definition explanation rather than a substantive risk difference. Cross-domain SHAP analysis identifies four instrument-independent CVD risk factors and two major portability failures.

**Conclusions:** EXHEART combines three practices that population-scale cardiovascular classifiers usually apply in isolation: leakage-free training with calibrated probabilities, a test of whether the model’s explanations are stable, and a fairness audit that examines intersecting subgroups rather than single attributes. Bringing them together proved worthwhile. The intersectional audit revealed disparities that single-attribute auditing missed, and the cross-instrument comparison indicated that much of the sex gap reflects how the outcome is measured in survey data rather than a substantive difference in risk. The temporal transport findings indicate that deployed BRFSS models warrant periodic monitoring and retraining to maintain clinical utility. EXHEART is a retrospective methodological evaluation on public de-identified data; it is not validated for direct clinical decision-making, diagnosis, or treatment recommendation without prospective clinical validation.

## 1. Introduction

Cardiovascular disease is the leading cause of death worldwide, accounting for roughly 17.9 million deaths annually, or 32% of all global fatalities [1], yet much of it is preventable through early identification of modifiable risk factors. Population-scale telephone surveys such as the Behavioral Risk Factor Surveillance System (BRFSS), conducted annually by the U.S. Centers for Disease Control and Prevention, offer a cost-effective alternative to hospital-based screening: they collect self-reported behavioral, demographic, and clinical indicators from hundreds of thousands of adults each year without requiring laboratory measurements [2]. Machine learning models trained on BRFSS data have consequently attracted substantial attention for community-level cardiovascular disease classification.

Despite this interest, three recurring methodological problems undermine the reliability of published BRFSS-based heart disease models, and none has been addressed jointly in the existing literature.

The first problem is data leakage through synthetic oversampling. BRFSS heart disease datasets exhibit severe class imbalance, approximately 9% positive prevalence in the 2015 cohort, leading many studies to apply synthetic minority oversampling (SMOTE or its variants) before splitting the data into training and test sets. This practice contaminates the validation procedure, inflating reported AUC and producing models whose apparent performance does not replicate in deployment [3,4]. A leakage-free pipeline that handles class imbalance exclusively through cost-sensitive training is therefore a prerequisite for credible performance claims.

The second problem is the qualitative treatment of explainability. Post-hoc explanation methods such as SHAP [5] and LIME [6] are widely deployed in clinical ML to communicate model reasoning to practitioners, yet the degree to which these two methods agree on which features matter, and under what conditions they diverge, is almost never quantified. Studies comparing SHAP and LIME on BRFSS data invariably do so through side-by-side bar charts, leaving clinicians with no quantitative basis on which to judge how often, and for which patients, the two methods would give contradictory guidance [7,8]. Without a formal consistency framework, clinical explainability remains aspirational rather than verified.

The third problem is the absence of demographic fairness evaluation at the operating-point level. Standard machine learning fairness audits are rarely applied to cardiovascular prediction models at the threshold used for clinical screening, and when they are, they examine only single protected attributes in isolation [9]. In practice, a young low-income woman faces compounding disadvantages that a sex-only or age-only audit cannot detect [10]. Moreover, no published study has examined whether BRFSS-derived fairness disparities are stable across survey years or across data collection instruments, a critical question for any model intended for sustained deployment.

This paper presents EXHEART, a fairness-aware explainable pipeline that addresses all three problems simultaneously on three complementary datasets. The primary dataset is the BRFSS 2015 Heart Disease Health Indicators subset (n = 253,680). To test temporal generalisability, we apply the trained model directly to the BRFSS 2020 survey (n = 319,795) without retraining (a feature-mismatch temporal transport stress test that quantifies degradation under combined temporal drift and feature mismatch), and then retrain independently on the 2020 cohort to measure recovery. To test cross-domain generalisability, we apply the same pipeline to the Cardiovascular Disease clinical examination dataset (n = 68,730), which provides objectively measured blood pressure, cholesterol, and glucose values in place of self-reported survey responses.

Our methodology delivers five measurable contributions. First, we construct a leakage-free stacked ensemble of four base learners, trained with class-weighted loss and combined via out-of-fold logistic regression stacking with Platt scaling, achieving AUC-ROC = 0.850 and a 96% ECE reduction on BRFSS 2015 (C1). Second, we report AUC, Brier score, ECE, and decision-curve net benefit at both overall and subgroup level across all three datasets, treating subgroup calibration gaps as an independent fairness dimension (C2). Third, we introduce a quantitative SHAP–LIME consistency framework using Kendall-τ rank correlation, Spearman-ρ, and Jaccard overlap at top-k, revealing a cross-domain inversion in top-k overlap: substantial top-3 divergence on survey data (Jaccard@3 = 0.200) but perfect top-3 agreement on clinical data (Jaccard@3 = 1.000, τ = 0.485, p = 0.031) (C3). Fourth, we conduct subgroup-stratified SHAP interaction analysis across sex, age, income, and race/ethnicity, demonstrating that pairwise interaction patterns differ substantially by demographic group (C4). Fifth, we introduce cross-instrument fairness-disparity attribution, an empirical diagnostic that tests whether a subgroup disparity is more consistent with a measurement-induced or a substantive explanation by re-validating it on an instrument that measures the same construct objectively, applied within a full intersectional fairness audit. The audit reveals Sex × Age disparities up to 5.2× the single-attribute gap among adequately-powered cells, and the attribution test indicates the female under-detection disparity is instrument-dependent rather than a substantive difference in risk, because it is not reproduced on clinical data (C5).

The remainder of the paper covers related work (Section 2), the datasets and methodology (Sections 3 and 4), per-dataset and cross-dataset results (Sections 5 and 6), and a discussion of clinical and methodological implications (Sections 7 and 8).

## 2. Related Work

### 2.1. BRFSS-Based Cardiovascular Disease Prediction

Machine learning approaches to BRFSS-based heart disease prediction have proliferated since the public availability of the Kaggle-derived 2015 indicator dataset [2]. Deng et al. [11] trained a LightGBM classifier on BRFSS 2015, achieving an AUROC of 0.811 with external validation on Framingham and Z-Alizadeh Sani cohorts, and applied SHAP to identify age, smoking, diabetes, hypertension, and high cholesterol as primary contributors. Tompra et al. [12] applied SMOTE-ENN oversampling with CatBoost on BRFSS 2021, reporting 88% recall and 82% AUC for high-risk patient identification. Ara et al. [13] compared seven machine learning classifiers on BRFSS 2020 with SMOTE balancing and three feature-selection schemes. While these studies demonstrate the feasibility of BRFSS-scale CVD prediction, none addresses data leakage through pre-split oversampling, none quantifies SHAP–LIME agreement, and none conducts a demographic fairness audit at the clinical operating threshold. EXHEART directly addresses all three limitations on three datasets.

### 2.2. Data Leakage in Machine Learning

Data leakage, broadly defined as the contamination of validation sets with information derived from the full dataset, is a pervasive source of over-optimistic performance claims in machine learning research. Kapoor and Narayanan [3] surveyed 17 fields and identified 294 affected papers, presenting a taxonomy of eight leakage subtypes. In the cardiovascular ML literature, the specific form of leakage introduced by applying synthetic oversampling before a train–test split has received renewed attention. Eltawil et al. [4] published a formal commentary demonstrating that SMOTE-ENN applied to the full cardiovascular dataset before a 70:30 split constitutes textbook leakage, with the corresponding authors acknowledging the methodological concern. EXHEART avoids this pitfall by applying class-weighted loss functions exclusively to training data, with no synthetic samples generated at any stage of the pipeline.

### 2.3. SHAP and LIME in Clinical Machine Learning

SHAP [5] and LIME [6] are the dominant post-hoc explanation methods in clinical ML. SHAP assigns each feature a value representing its marginal contribution to the model output based on Shapley values from cooperative game theory, whereas LIME approximates model behaviour locally with a linear surrogate trained on perturbed instances. A systematic review by Vimbi et al. [7] identified 23 studies applying LIME and SHAP to Alzheimer’s disease detection, noting that the methods frequently diverge in their rankings despite agreeing on the broad importance of well-established clinical features. An evaluation on UK primary-care lung cancer data [8] measured SHAP–LIME consistency explicitly using Jaccard similarity and rank agreement, finding that consistency decreases under class imbalance, directly relevant to BRFSS’s 9:1 ratio. A meta-analysis of medical imaging models [14] found LIME achieving higher fidelity than SHAP, contrasting with tabular results; this suggests method agreement is modality-dependent. EXHEART addresses this gap by introducing a quantitative consistency framework (Kendall-τ, Spearman-ρ, and Jaccard@k) applied across three datasets to test whether agreement patterns are instrument-dependent.

### 2.4. Fairness Auditing in Clinical AI

Hardt et al. [9] formalised equal opportunity as the requirement that a classifier’s true positive rate be equal across protected groups, establishing the theoretical basis for the TPR-parity audit we conduct in this paper. Obermeyer et al. [15] demonstrated that a widely deployed healthcare algorithm exhibited systematic under-referral of Black patients, with disparity-adjusted re-analysis raising the proportion identified for extra care from 17.7% to 46.5%. Seyyed-Kalantari et al. [16] showed that AI-based chest radiograph models exhibit underdiagnosis bias with TPR disparities exceeding 18 percentage points for under-served demographic groups, with intersectional subgroups facing compounded disadvantage. The importance of intersectional analysis, evaluating subgroup combinations rather than individual attributes, was established by Buolamwini and Gebru [10], whose Gender Shades study revealed error rates up to 34.7% for darker-skinned females compared with 0.8% for lighter-skinned males in commercial systems. Xu et al. [17] provided a clinical review formalising demographic parity, equalized odds, and equal opportunity for computational medicine. To our knowledge, no published BRFSS cardiovascular prediction study has applied intersectional fairness analysis or tested whether disparities are stable across survey years or data collection instruments; EXHEART addresses both gaps.

### 2.5. Probability Calibration

Probability calibration, the alignment between predicted probabilities and empirical event frequencies, is essential for clinical decision support, where output scores are used directly for risk-stratified intervention [18]. Van Calster et al. [18] established calibration as the “Achilles heel” of predictive analytics and articulated the calibration hierarchy from mean to flexible calibration. Niculescu-Mizil and Caruana [19] demonstrated that Platt scaling (logistic regression on model outputs) and isotonic regression are the two most effective post-hoc calibration methods, with Platt preferred for small calibration sets and isotonic for larger ones. A clinical tutorial by Huang et al. [20] operationalised ECE, Brier score, and Spiegelhalter’s Z for clinical prediction model evaluation. EXHEART applies Platt scaling to the stacked ensemble output and reports both pre- and post-calibration ECE, demonstrating a 96% ECE reduction on BRFSS survey data while showing that Platt marginally increases ECE on the balanced clinical dataset, a context-dependent result not previously reported for CVD prediction.

### 2.6. Temporal Model Drift

Temporal dataset shift, changes in the joint distribution of features and outcome over time, is a well-documented cause of clinical ML degradation in deployment. Guo et al. [21] benchmarked domain generalisation and adaptation methods against temporal shift on MIMIC-IV across a 12-year period, finding that naive retraining consistently outperformed sophisticated domain adaptation techniques. Nestor et al. [22] quantified feature robustness under non-stationarity in MIMIC-III, identifying clinically meaningful performance degradation driven by EHR system transitions rather than true population change. Alghamdi et al. [23] assessed data drift in sepsis prediction models, distinguishing concept drift from covariate shift. No BRFSS-based cardiovascular prediction study has, to our knowledge, quantified temporal drift by applying a frozen trained model directly to a later survey wave without retraining; EXHEART fills this gap with a five-year transport experiment and measures AUC, ECE, sensitivity, and fairness metric drift simultaneously.

### 2.7. Cross-Domain Generalisation

Cross-domain generalisability, the ability of a model trained on one data collection instrument to maintain performance on a different instrument, is distinct from temporal generalisation and remains understudied in cardiovascular ML. Zhang et al. [24] benchmarked domain generalisation methods on clinical imaging and structured time series, finding that out-of-distribution performance frequently fails to surpass standard empirical risk minimisation. Maadi et al. [25] demonstrated site-specific performance clustering across four NHS sites for COVID-19 screening, with AUC dropping by up to 0.12 when applied to an external site. Bhatt et al. [26] proposed a unified framework distinguishing internal and external validation, temporal validation, and cross-domain validation as progressively demanding test conditions. EXHEART applies this hierarchy by validating on a clinical examination dataset that differs from BRFSS not only in population but in measurement modality, replacing self-reported binary flags with objectively measured continuous values for blood pressure, cholesterol, and glucose.

### 2.8. Fairness-Gap Attribution and Measurement Bias

Attributing a fairness gap to its sources is an established methodological direction. Plečko and Bareinboim [34] decompose the total disparity between groups into direct, indirect, and spurious causal components, and Zhang et al. [35] attribute a gap to fairness-aware causal paths running from the protected attribute to the prediction. Schrouff et al. [36] diagnose why fairness transfers or fails across distribution shift by separating demographic, covariate, and label contributions. Closest in spirit to the present work, a recent imaging study shows that a technical acquisition parameter (radiograph view type) explains the majority of an apparent demographic performance gap in chest X-ray models, far exceeding the contribution of sex or age [37]. The underlying phenomenon also has a statistical precedent: Mikhaeil et al. [38] formalise how a proxy or self-reported label, when its error correlates with predictors, induces group-correlated prediction disparities. Within cardiovascular surveillance specifically, Thakkar et al. [39] show that BRFSS reports a higher heart-attack prevalence than the examination-based NHANES for the same outcome, with race-varying differences, confirming that the data collection instrument itself shifts measured disparities. EXHEART differs from this body of work in a specific combination: it uses operating-point fairness metrics to compare the same disease outcome under a self-report instrument and a clinical-examination instrument, and associates the resulting gap with the outcome-measurement channel. Where the imaging work isolates an input-acquisition artefact, EXHEART provides diagnostic evidence pointing to an outcome-labelling channel; we position our test as an empirical, cross-instrument instantiation of fairness-gap attribution rather than a new decomposition method.

## 3. Datasets

### 3.1. BRFSS 2015 (Primary Dataset)

The primary dataset is derived from the BRFSS 2015 annual telephone survey conducted by the CDC across all 50 U.S. states, the District of Columbia, and participating territories [2]. We use the publicly available cleaned subset (n = 253,680) prepared by Teboul [27] and hosted on Kaggle, which retains 21 binary and ordinal features alongside a binary cardiovascular disease indicator (HeartDiseaseorAttack). The positive class prevalence is 9.4%, yielding a 9.6:1 class imbalance ratio. Features capture four domains: demographic variables (Sex, Age in 13 bands, Education, Income), clinical indicators (HighBP, HighChol, CholCheck, Stroke, Diabetes), lifestyle behaviors (BMI, Smoker, AlcoholDrinking, PhysActivity, Fruits, Veggies), and health status (GenHlth, MentHlth, PhysHlth, DiffWalk, AnyHealthcare, NoDocbcCost). All features are self-reported. Demographically reweighted evaluation reweights the test set by age and sex to approximate the U.S. adult population distribution; this is a post-hoc demographic adjustment, not the formal CDC BRFSS complex survey design weighting.

### 3.2. BRFSS 2020 (Temporal Validation)

The BRFSS 2020 dataset is sourced from the cleaned version published by Kumar and Pande [28] on Zenodo (DOI: 10.5281/zenodo.15364962), derived from the CDC’s 401,958-row 2020 survey. After cleaning, 319,795 records are retained with 18 features, including HeartDisease (binary target), BMI, Smoking, AlcoholDrinking, Stroke, PhysicalHealth, MentalHealth, DiffWalking, Sex, AgeCategory (14 bands), Race (six categories: American Indian/Alaskan Native, Asian, Black, Hispanic, Other, White), Diabetic, PhysicalActivity, GenHealth, SleepTime, Asthma, KidneyDisease, and SkinCancer. The positive class prevalence is 8.6%. Critically, BRFSS 2020 includes a race/ethnicity indicator absent from the 2015 dataset, enabling a racial fairness audit not possible with the primary dataset. Eleven features have conceptual equivalents across the two survey waves; ten features present in 2015 (including HighBP and HighChol) have no equivalent in 2020 and are imputed with 2015 training-set medians for the temporal transport experiment.

### 3.3. Cardiovascular Disease Dataset (Cross-Domain Validation)

The Cardiovascular Disease dataset [29], sourced from Kaggle, contains 70,000 clinical examination records collected at the moment of medical assessment. After removing physiologically implausible values (systolic BP < 60 or > 250 mmHg, diastolic BP < 40 or > 160 mmHg, height outside 100–220 cm, weight outside 30–200 kg), 68,730 records are retained. The target variable (cardio) indicates cardiovascular disease presence, with near-balanced class distribution (49.5% positive). Features include age (converted from days to years; range 29.6–64.9), gender (1 = female, 2 = male), height, weight, systolic BP (ap_hi), diastolic BP (ap_lo), cholesterol (1 = normal, 2 = above normal, 3 = well above normal), glucose (same scale), smoking status, alcohol consumption, physical activity, and derived BMI. Unlike BRFSS, all measurements are objective clinical recordings rather than self-reported responses. The dataset contains no race/ethnicity variable and no patients under 30 or over 65. The near-balanced prevalence (49.5%) means the BRFSS operational threshold (pt = 0.12) is inappropriate; all Cardio results use pt = 0.50 as the primary threshold, with pt = 0.12 reported solely for direct cross-dataset comparison.

**Table 1.** Characteristics of the three experimental datasets.

| Property | BRFSS 2015 | BRFSS 2020 | Cardio Dataset |
| --- | --- | --- | --- |
| Source | CDC BRFSS / Kaggle | CDC BRFSS / Zenodo | Kaggle (Ulianova) |
| Total records (cleaned) | 253,680 | 319,795 | 68,730 |
| Test set size | 50,736 | 63,959 (retrain); 319,795 | 13,746 |
|  |  | (transport) |  |
| **Feature count** | 21 | 18 | 12 |
| **Positive prevalence** | 9.4% | 8.6% | 49.5% |
| **Age range** | 18–80+ yrs | 18–80+ yrs | 29.6–64.9 yrs |
| **Race/ethnicity** | Not available | 6 categories | Not available |
| **Measurement type** | Self-reported survey | Self-reported survey | Clinical exam |
| **Primary threshold** | pt = 0.12 | pt = 0.12 | pt = 0.50 |
| **Class imbalance ratio** | 9.6:1 | 10.7:1 | 1.02:1 |

BRFSS = Behavioral Risk Factor Surveillance System. pt = operating threshold.

## 4. Methodology

### 4.1. Leakage-Free Preprocessing

All preprocessing is applied strictly after the train–test split, with no information from the test set used in any transformation. Categorical features in BRFSS 2020 (encoded as string labels) are label-encoded using a LabelEncoder fitted on the training set. This integer mapping is exact for binary indicators, which constitute most of the 2020 categorical features; for the nominal Race variable, which has no natural order, label encoding imposes an arbitrary ordinal scale, a recognised limitation; we therefore re-ran the retrained pipeline with a one-hot encoding of race and confirmed that the race-feature conclusion is directionally unchanged (reported in the Discussion), so the finding does not depend on the label-encoding of race. Alphabetical label encoding preserves the AgeCategory order but not the GenHealth ordinal scale, a minor limitation that the tree base learners, which can split a feature repeatedly, largely absorb. A StandardScaler for the MLP base learner is also fitted exclusively on training data and applied to both training and test partitions at inference time. No synthetic oversampling is applied at any stage. Class imbalance is addressed through sample weighting in the loss function of each base learner: XGBoost uses scale_pos_weight equal to the ratio of negative to positive training samples; LightGBM and Random Forest use class_weight=“balanced”; the MLP uses class-weighted binary crossentropy. All datasets are split 80%/20% with stratification on the target variable and random seed 42.

### 4.2. Base Learners

Four base learners are trained independently on the training set. XGBoost [30] is configured with 300 estimators, max_depth = 6, learning_rate = 0.05, and scale_pos_weight = 9.6 (BRFSS 2015). LightGBM [31] uses 300 estimators, max_depth = 6, learning_rate = 0.05, and class_weight = “balanced”. Random Forest uses 200 trees, max_depth = 10, and class_weight = “balanced”. The MLP consists of Dense(256, ReLU) → BatchNormalization → Dropout(0.3) → Dense(128, ReLU) → BatchNormalization → Dropout(0.3) → Dense(64, ReLU) → Dropout(0.2) → Dense(1, Sigmoid), trained with Adam (lr = 1 × 10⁻³) with early stopping on validation AUC (patience = 5) and learning-rate reduction on plateau (factor = 0.5, patience = 3). Batch size is 512 for all BRFSS datasets and the Cardio dataset.

### 4.3. Stacked Ensemble

A second-level Logistic Regression meta-learner is trained on out-of-fold (OOF) predictions generated by 5-fold stratified cross-validation. For each fold, the four base learners are re-trained on the in-fold training partition, and their predicted positive-class probabilities on the held-out validation partition constitute four-column OOF features. After all five folds, the meta-learner is fitted on the concatenated OOF predictions using the true labels. For test-set inference, the full base learners (trained on the entire training set) generate four probability columns, which are passed to the fitted meta-learner. Meta-learner coefficients are reported for each dataset to quantify the relative contribution of each base learner and to test for dataset-specific weighting patterns.

### 4.4. Calibration

Raw stacked ensemble probabilities are calibrated using manual Platt scaling: a univariate Logistic Regression is fitted on a 20% calibration hold-out of the training set (stratified by target) using the raw stack probabilities as the single input feature. The fitted Platt scaler is applied to test-set stack probabilities before all threshold-dependent evaluations. Pre- and post-calibration Expected Calibration Error (ECE) is computed over 10 equal-width bins. The Brier score is computed on post-calibration probabilities.

### 4.5. Evaluation Metrics and Operating Threshold

Primary discrimination metrics are AUC-ROC and AUPRC (area under the precision-recall curve). Calibration is assessed by ECE (pre- and post-Platt) and Brier score. At the clinical operating threshold (pt = 0.12 for BRFSS; pt = 0.50 for Cardio), we report sensitivity, specificity, and the confusion matrix. Decision-curve analysis (DCA) computes the net benefit of the model against treat-all and treat-none strategies across a range of threshold probabilities (0.01–0.50). Demographically reweighted AUC and Brier score for BRFSS 2015 are computed by reweighting the test set to the U.S. adult population age and sex distribution, a post-hoc demographic adjustment rather than formal BRFSS survey weighting.

### 4.6. SHAP Analysis

Global feature importance is computed using TreeSHAP [5] on the XGBoost base learner over 2,000 randomly sampled test instances. For consistency, both the SHAP and LIME agreement analyses explain the XGBoost base learner; stacked-ensemble explanations are left for future model-agnostic analysis. Pairwise SHAP interaction values are computed on 500 test instances. All interaction heatmaps report mean absolute interaction magnitude with main effects zeroed on the diagonal. Subgroup-stratified SHAP analysis partitions the test set by Sex, Age band, and Income band, computing mean absolute SHAP per feature per stratum to identify differential interaction patterns across demographic groups.

### 4.7. LIME Analysis and SHAP–LIME Consistency

LIME explanations [6] are generated for 200 randomly sampled test instances using LimeTabularExplainer with num_features equal to the full feature set and 500 perturbation samples per instance. Global LIME importance is the mean absolute weight across all 200 instances. SHAP–LIME consistency is quantified by Kendall-τ rank correlation, Spearman-ρ rank correlation, and Jaccard overlap at k = 3 and k = 5 (Jaccard@k = |SHAP_top_k ∩ LIME_top_k| / |SHAP_top_k ∪ LIME_top_k|). P-values are computed using the exact distribution for Kendall-τ and the normal approximation for Spearman-ρ. The analysis is repeated on the Cardio dataset to test whether consistency patterns are instrument-dependent.

### 4.8. Fairness Audit and Mitigation

Fairness is evaluated at the clinical operating threshold. For each protected attribute (Sex, Age, Income on BRFSS 2015; Sex, Age, Race/Ethnicity on BRFSS 2020; Gender, Age on Cardio), we compute the true positive rate (TPR), selection rate, and prevalence per group. The primary fairness metric is the TPR gap (max TPR − min TPR across groups), operationalising equal opportunity [9]. Subgroup ECE is computed per group as an additional fairness dimension: a well-calibrated model overall may exhibit systematic miscalibration for minority subgroups. Intersectional fairness is evaluated for Sex × Age, Sex × Income, and Age × Income cells on BRFSS 2015, with cells containing fewer than 30 positive cases excluded, since the true positive rate is unstable when a cell has very few positives. Mitigation applies threshold shifting per group, sweeping the mitigation strength parameter from 0 to 1 in 20 steps. A frontier of Sex TPR gap versus overall balanced accuracy is generated across the sweep; because mitigation operates purely by per-group threshold shifting, AUC is invariant by construction and serves only as a sanity check, with balanced accuracy and net benefit used as the threshold-dependent utility measures.

To attribute observed disparities to their source, we apply cross-instrument disparity attribution. For each protected-attribute disparity measured on the survey instrument, we re-evaluate the equivalent disparity on the clinical examination dataset, which captures the same clinical construct through objective measurement rather than self-report. A disparity shows diagnostic evidence of being measurement-induced when the clinical TPR gap collapses toward parity and the attribution of the responsible feature falls correspondingly in the SHAP ranking, and of being substantive when it persists at a comparable magnitude. This diagnostic evidence informs the choice of remedy: measurement-induced disparities call for instrument redesign or instrument-aware deployment, whereas substantive disparities require algorithmic mitigation such as the threshold shifting described above. The validity of the test depends on construct matching, since both instruments must measure the same underlying clinical quantity, a condition that holds for sex, age, blood pressure, and cholesterol across the survey and examination datasets used here. This is an empirical, cross-instrument attribution: it compares operating-point fairness metrics across datasets rather than identifying a causal effect within a single structural model, and we therefore make no claim of causal identifiability.

For the temporal transport experiment, BRFSS 2015 model parameters (XGBoost, LightGBM, Random Forest, meta-learner, and Platt scaler) are frozen and applied directly to BRFSS 2020. Missing 2015 features are imputed with training-set medians. Performance drift, ECE drift, sensitivity drift, and fairness drift are computed as signed differences between 2015 test-set metrics and 2020 transport metrics.

## 5. Results

### 5.1. BRFSS 2015: Performance and Calibration

On the BRFSS 2015 test set (n = 50,736; 9.4% positive), the stacked ensemble achieves AUC-ROC = 0.850, AUPRC = 0.371, and Brier score = 0.071 (Table 2). The demographically reweighted AUC is 0.860, marginally higher than the unweighted estimate, indicating that BRFSS oversampling of older high-risk adults slightly deflates apparent discrimination. At the clinical screening threshold (pt = 0.12), sensitivity is 0.776 and specificity is 0.769. The MLP is the strongest individual base learner (standalone AUC = 0.850) and receives the highest meta-learner coefficient (2.998), compared with XGBoost (0.092), LightGBM (0.882), and Random Forest (1.572), indicating that the neural network adds the most unique signal not captured by the tree ensemble.

**Table 2.** Model performance metrics across all experimental conditions.

| Metric | BRFSS 2015 | 2020 Transport | 2020 Retrained | Cardio ($p_t=0.5$) |
| --- | --- | --- | --- | --- |
| AUC-ROC | 0.850 | 0.781 | 0.840 | 0.807 |
| AUPRC | 0.371 | 0.227 | 0.353 | 0.788 |
| Brier Score | 0.071 | 0.076 | 0.066 | 0.180 |
| ECE (pre-calibration) | 0.256 | — | 0.267 | 0.018 |
| ECE (post-calibration) | 0.011 | 0.050 | 0.012 | 0.027 |
| Sensitivity | 0.776 | 0.267 | 0.731 | 0.687 |
| Specificity | 0.769 | 0.931 | 0.784 | 0.785 |
| AUC (demographically reweighted) | 0.860 | — | — | — |

AUC = area under ROC curve; AUPRC = area under precision-recall curve; ECE = expected calibration error; — = not applicable.

**Fig. 1.**
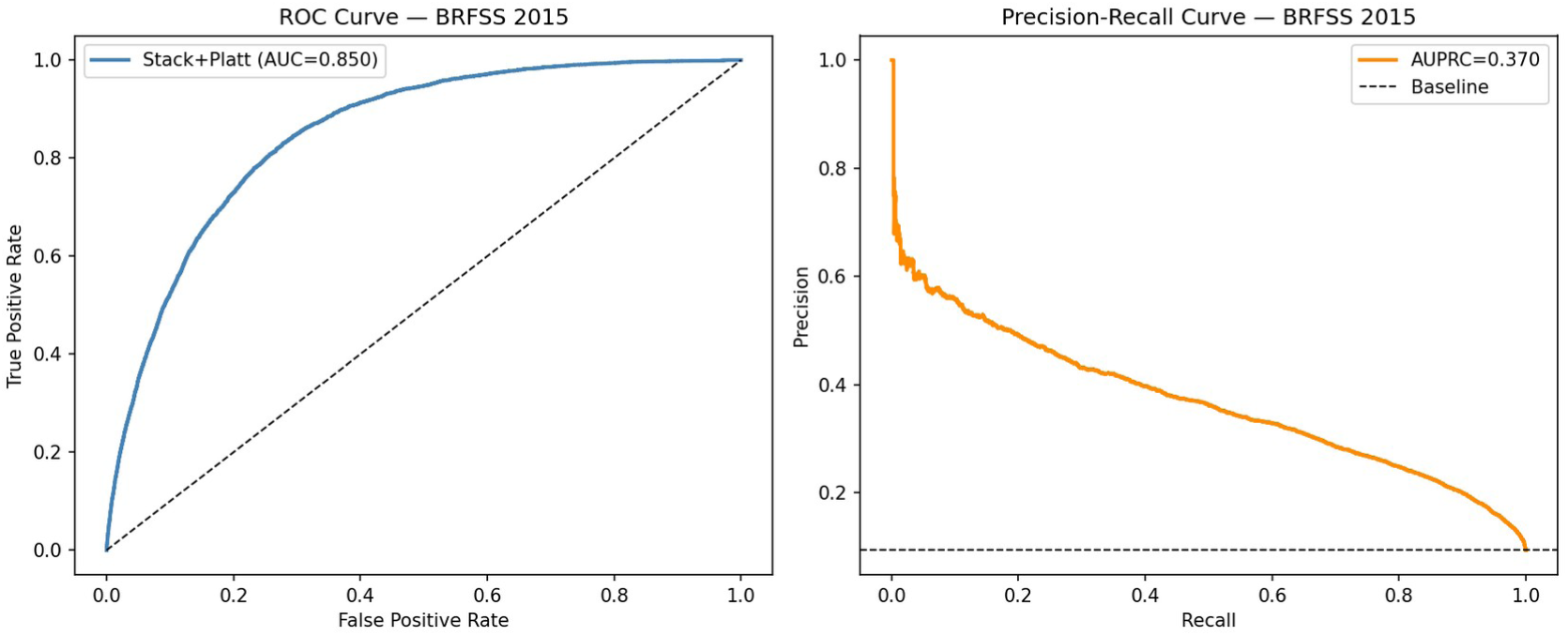
ROC curve (AUC = 0.850) and Precision-Recall curve (AUPRC = 0.371) for the EXHEART stacked ensemble on the BRFSS 2015 test set (n = 50,736). Both curves demonstrate strong discriminative performance despite a 9.6:1 class imbalance.

Platt scaling reduces ECE from 0.256 to 0.011, a 96% reduction, demonstrating that the raw stacked probabilities are substantially miscalibrated prior to post-hoc adjustment. Decision-curve analysis confirms positive net benefit over treat-all and treat-none baselines across the threshold range 0.05–0.40, validating the clinical utility of the model for population-level screening.

**Fig. 2.**
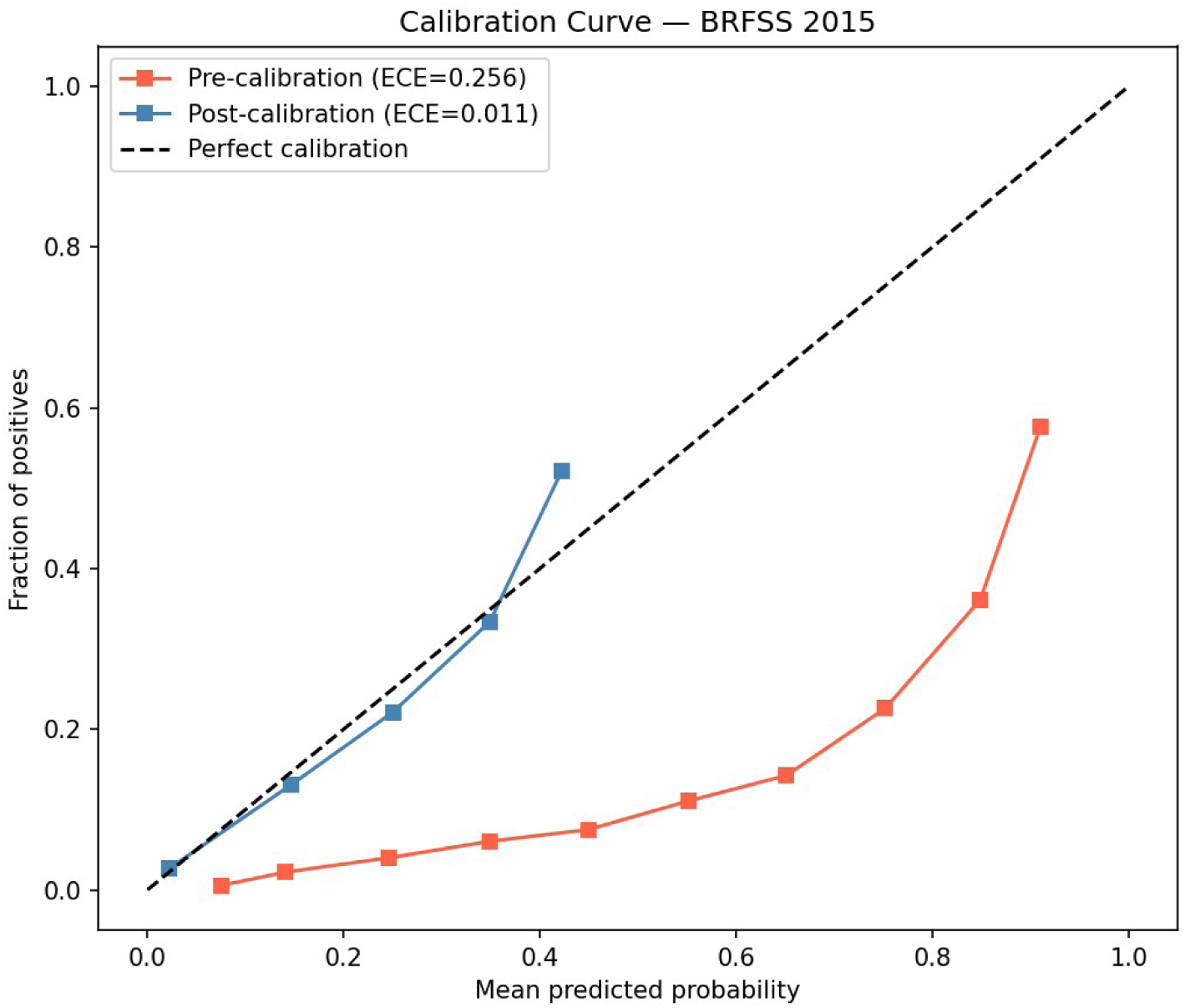
Calibration curves before (ECE = 0.256, red) and after (ECE = 0.011, blue) Platt scaling on BRFSS 2015. The post-calibration curve closely follows the diagonal, indicating good held-out calibration of the model-output probabilities on BRFSS 2015.

**Fig. 3.**
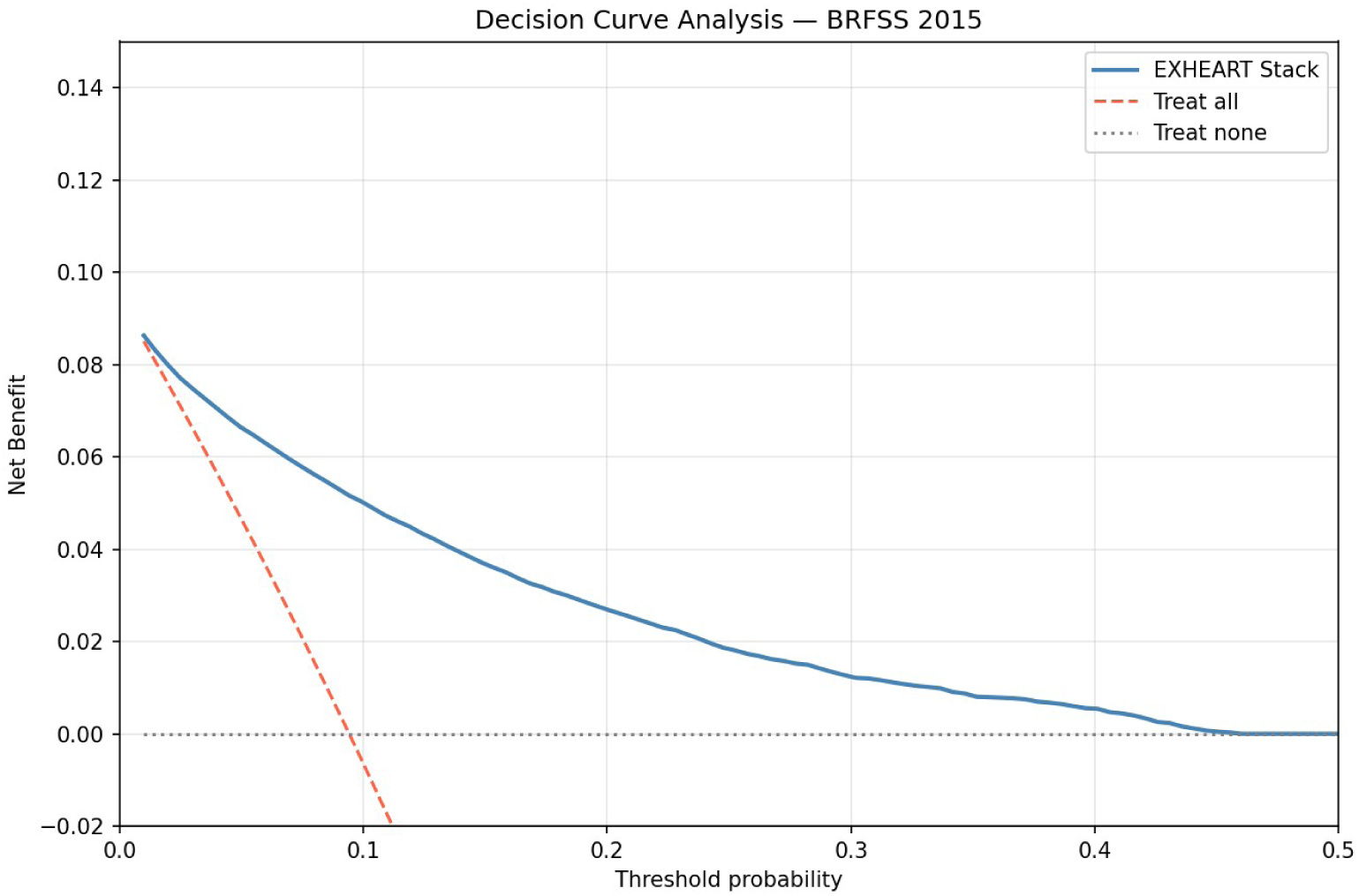
Decision Curve Analysis on BRFSS 2015. The EXHEART stack provides positive net benefit over treat-all (dashed red) and treat-none baselines across the clinically relevant threshold range 0.05–0.40.

**Fig. 4.**
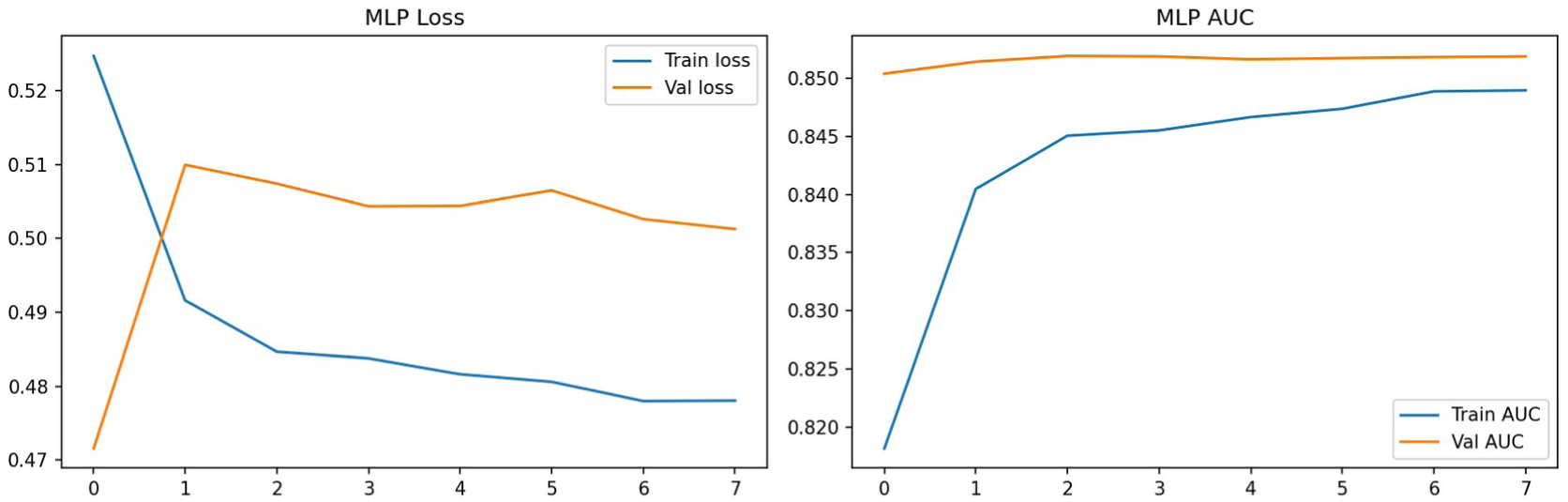
MLP training history on BRFSS 2015. Early stopping triggers at epoch 15. Validation AUC = 0.8517 with healthy convergence and no overfitting observed.

### 5.2. BRFSS 2015: SHAP and LIME Explainability

Age is the dominant predictor (mean |SHAP| = 0.764), followed by GenHlth (0.497), HighBP (0.474), Sex (0.355), and HighChol (0.322). The prominence of GenHlth, a subjective self-rating of overall health, above objective clinical indicators such as HighBP and HighChol is noteworthy and reflects the information value of patient-reported health perception in population survey models. Pairwise SHAP interaction analysis reveals the strongest interactions between Age × GenHlth and Age × HighBP, consistent with age-dependent hypertension risk.

Global LIME rankings show Stroke as the top-ranked feature, contrasting with its SHAP rank of 8 (mean | SHAP| = 0.128). This inversion indicates that Stroke has a highly localised, context-dependent effect: its globally moderate Shapley contribution masks a very strong local influence for the subset of patients for whom it is the most predictive feature. Quantitative consistency analysis yields Kendall-τ = 0.580 (p < 0.001) and Spearman-ρ = 0.818 (p < 0.001), indicating moderate-to-strong global rank agreement. However, Jaccard@3 = 0.200 (SHAP top-3: Age, GenHlth, HighBP; LIME top-3: Stroke, HighBP, DiffWalk), confirming a substantial divergence at the level most relevant for clinical communication.

**Fig. 5.**
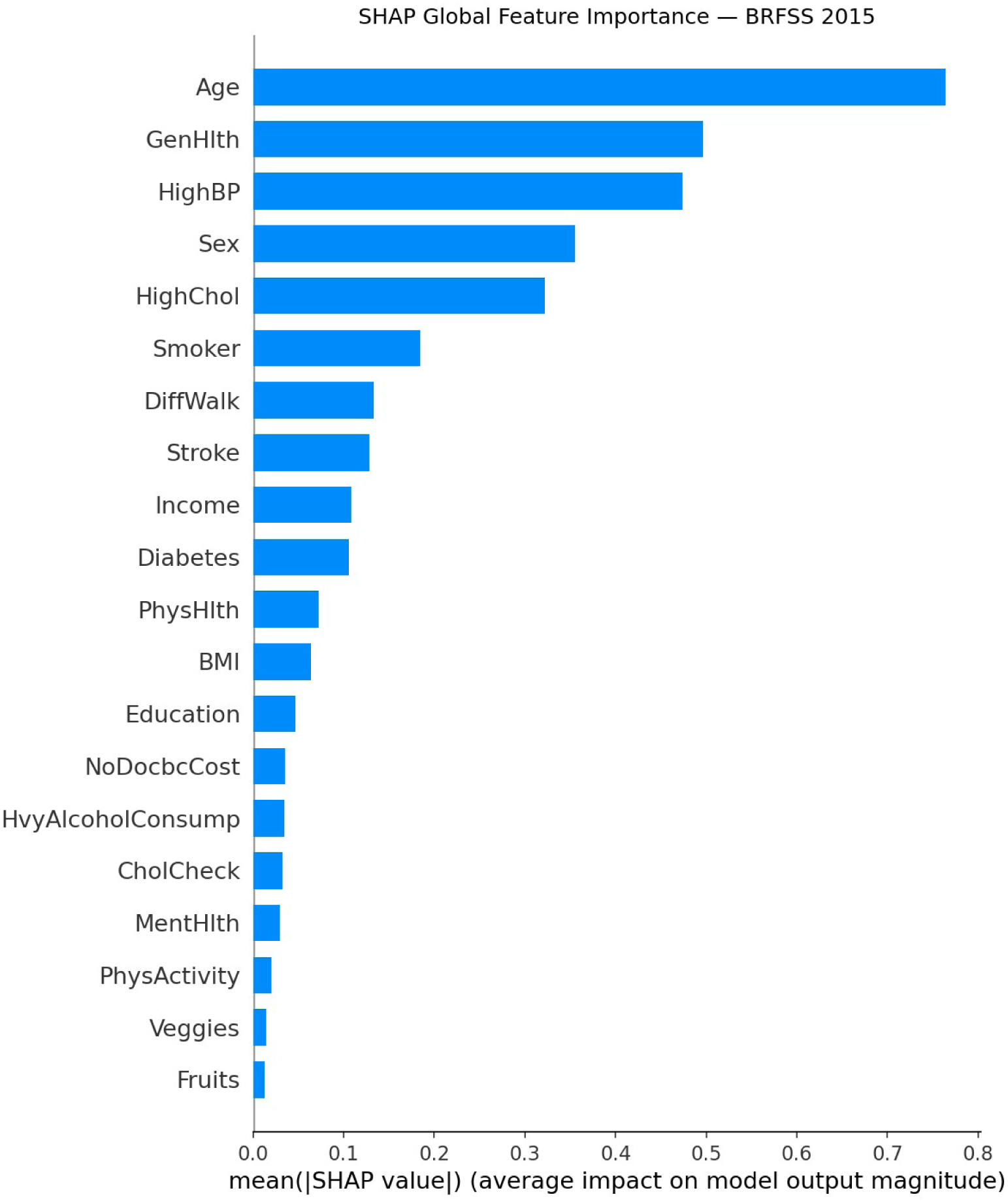
Global SHAP feature importance (mean |SHAP value|) on BRFSS 2015. Age dominates (0.764), followed by GenHlth (0.497) and HighBP (0.474). The prominence of GenHlth, a subjective self-rating, above objective clinical measurements is noteworthy.

**Fig. 6.**
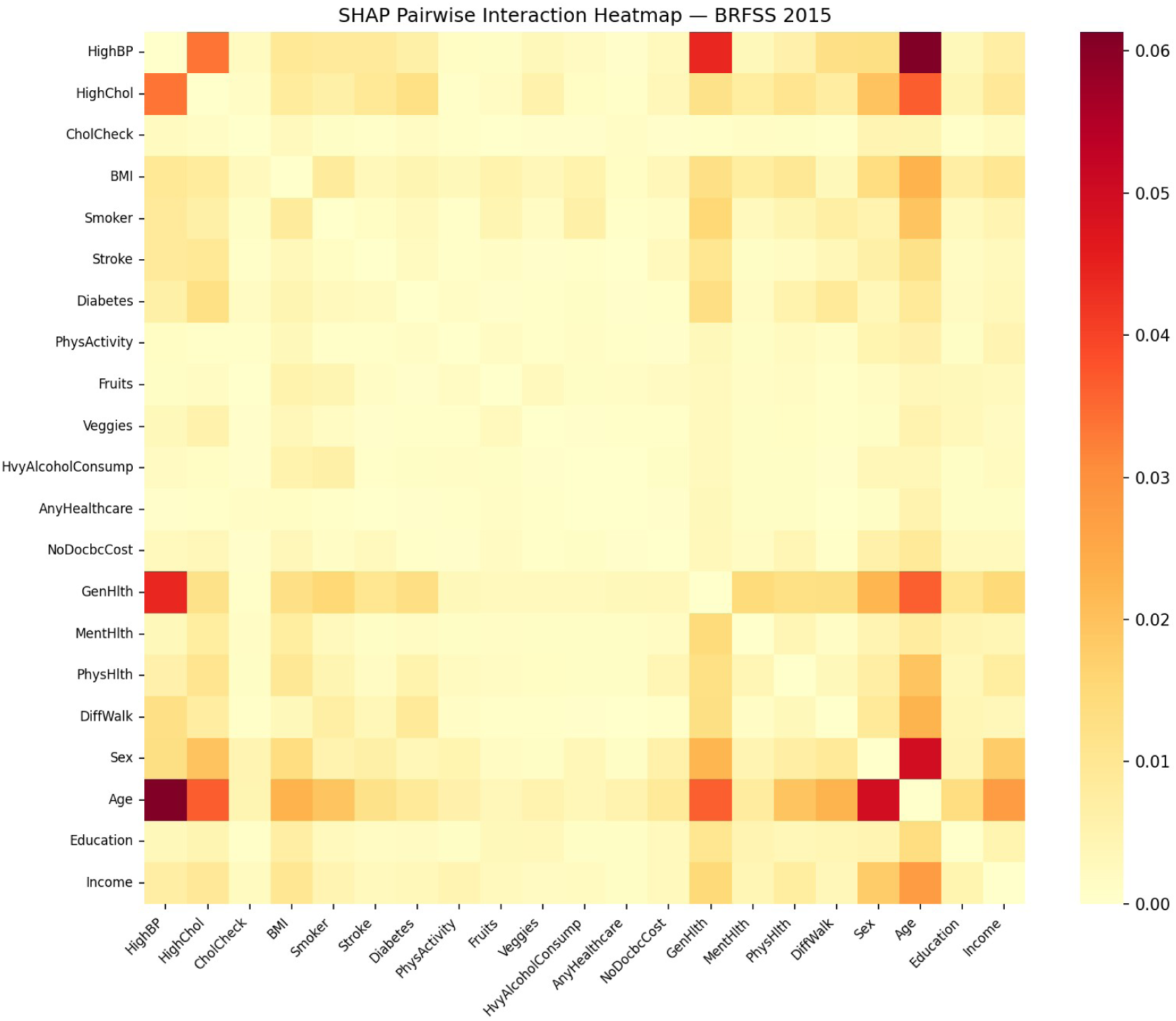
Pairwise SHAP interaction heatmap on BRFSS 2015 (n = 500 test instances). Warmer colours indicate stronger pairwise interaction magnitude. Age × GenHlth and Age × HighBP exhibit the strongest interactions, consistent with age-dependent CVD risk pathways.

**Fig. 7.**
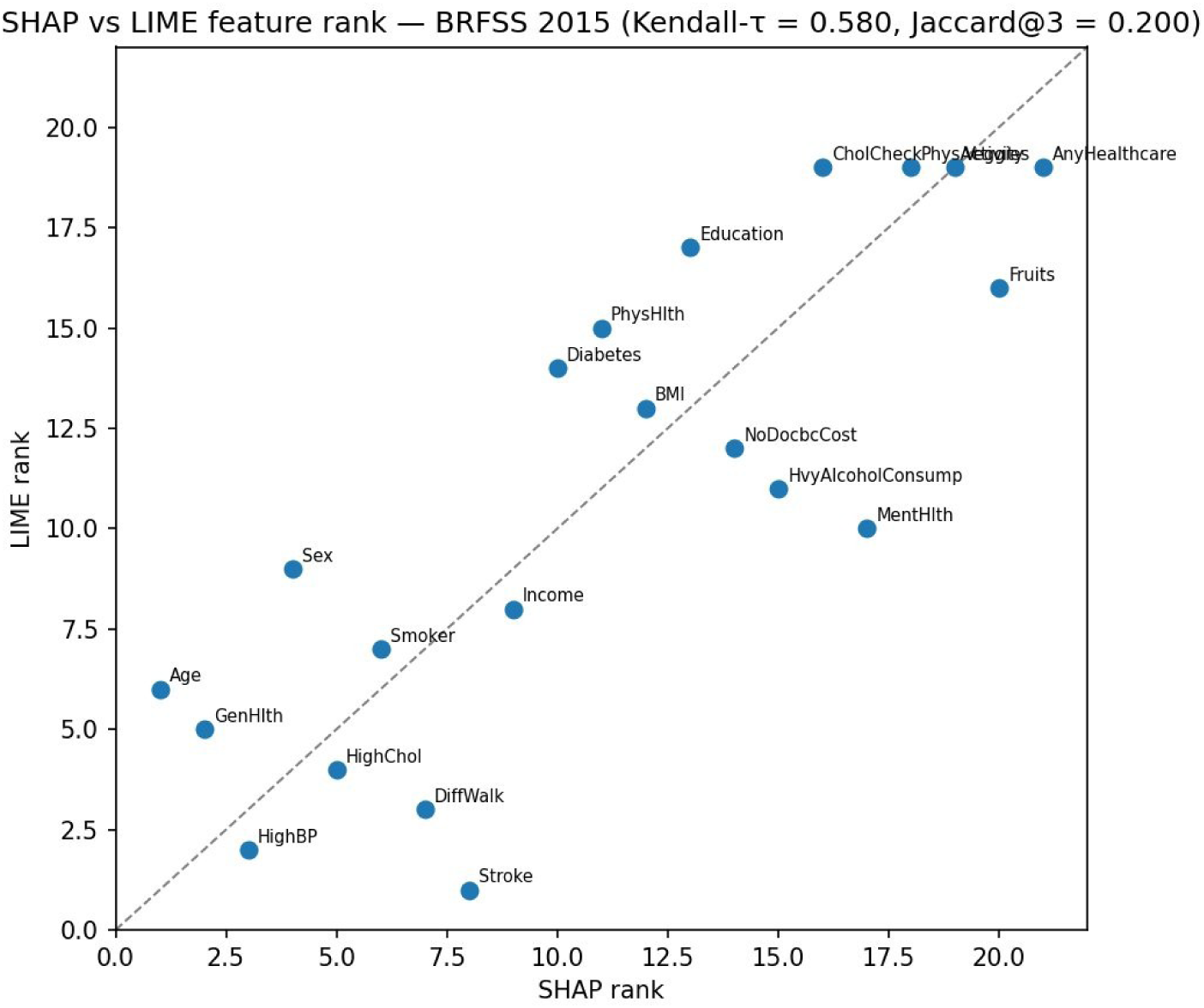
SHAP vs LIME feature rank scatter plot on BRFSS 2015 (Kendall-τ = 0.580, Jaccard@3 = 0.200). Stroke (SHAP rank 8, LIME rank 1) is the key outlier, indicating a context-dependent local effect not fully reflected in global SHAP attribution.

### 5.3. BRFSS 2015: Fairness Audit and Mitigation

At pt = 0.12, the Sex TPR gap is 0.124 (Female TPR = 0.705, Male TPR = 0.829). The Age TPR range spans 0.000 to 0.900: age group 2 (25–30 years) has TPR = 0.000, while age group 13 (80+ years) has TPR = 0.900. This near-complete failure on young adults is a direct consequence of Age being the dominant SHAP feature: the model cannot distinguish high-risk young patients from the low-prevalence majority. The Income TPR gap is 0.230, with an inverted gradient: TPR falls from 0.876 in lower-income groups to 0.646 in the highest-income group, reflecting that high-income patients fall below the screening threshold due to their lower prevalence.

Intersectional analysis reveals a maximum Sex × Age TPR gap of 0.649 (95% CI [0.545, 0.814]) among cells with at least 30 positive cases, 5.2× the Sex-only gap, with comparably large Age × Income disparities. Raw intersectional cells containing very few positives, such as the youngest female band with only two positive cases, produce extreme apparent gaps (a nominal 0.965) that are not statistically reliable and are excluded by this positive-count floor. These findings demonstrate that single-attribute fairness audits substantially understate true disparities when protected attributes interact. Subgroup ECE analysis shows that even after global Platt calibration, some demographic subgroups retain miscalibration gaps of up to 0.045, warranting group-specific calibration in high-stakes deployment.

**Fig. 8.**
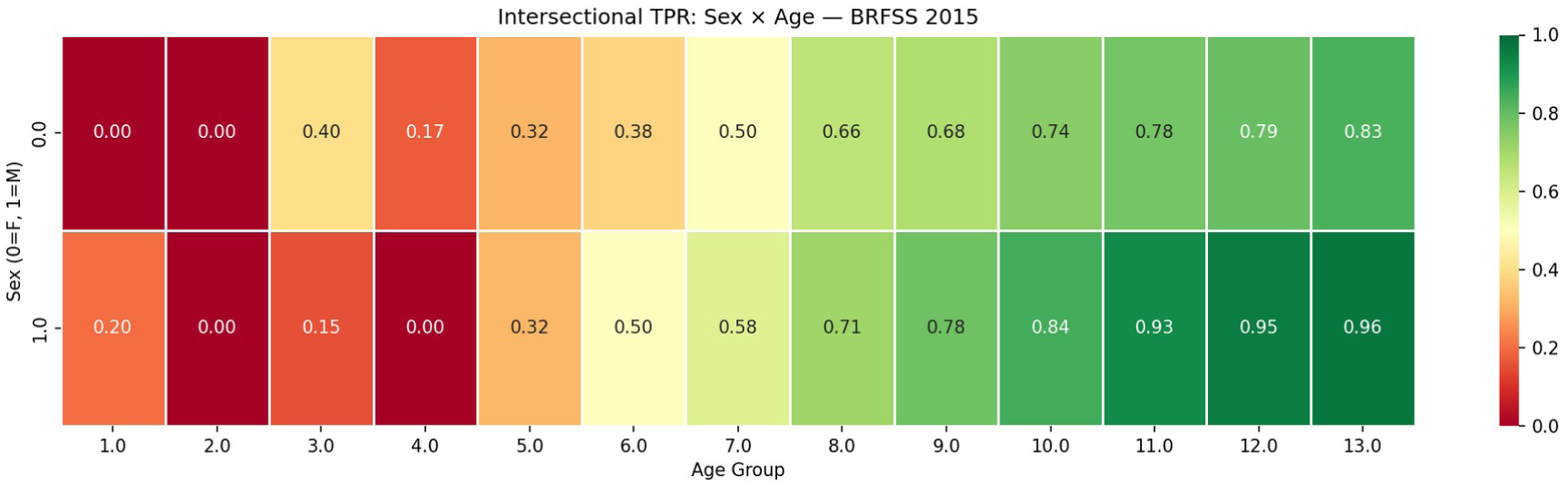
Intersectional TPR heatmap (Sex × Age) on BRFSS 2015. Young females (rows 0, columns 1–4) are the hardest-to-detect group. Among cells with at least 30 positive cases, the maximum Sex × Age TPR gap is 0.649 (95% CI [0.545, 0.814]), 5.2× the single-attribute Sex gap of 0.124; the lowest-TPR raw cells (youngest females) contain too few positives to estimate reliably and are not used for the headline gap.

Per-group threshold shifting reduces the Sex TPR gap from 0.124 to approximately 0.000 at full mitigation strength (s = 1.0), a near-complete elimination of the disparity. Because mitigation only moves per-group decision thresholds, AUC is invariant (0.850) and overall balanced accuracy is essentially unchanged (0.773 to 0.773); the cost manifests instead as a sensitivity-for-specificity rebalancing, with overall sensitivity rising from 0.776 to 0.829 while specificity falls from 0.769 to 0.716 and net benefit at the screening threshold falling only marginally (0.045 to 0.043). The frontier is smooth with no abrupt cliff edges, indicating that intermediate operating points are clinically achievable without catastrophic performance loss.

**Fig. 9.**
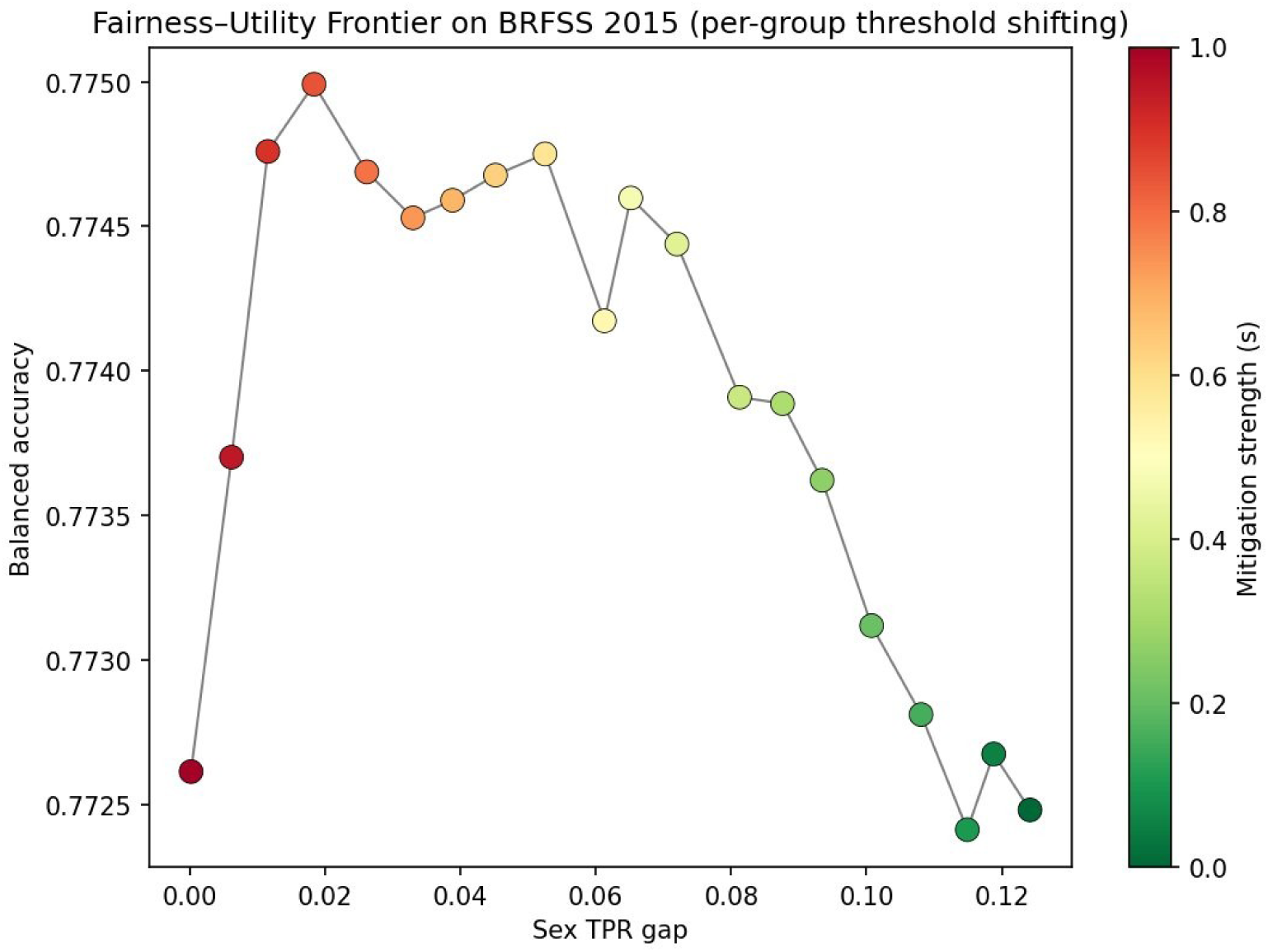
Fairness–utility frontier on BRFSS 2015 (per-group threshold shifting). Each point is one mitigation strength (green = no mitigation, red = full equalisation); the y-axis is overall balanced accuracy, a threshold-dependent utility. The Sex TPR gap reduces from 0.124 to approximately 0.000 while balanced accuracy stays within 0.772 to 0.775 and AUC is invariant under threshold shifting.

### 5.4. BRFSS 2020: Temporal Transport and Retraining

Applied to BRFSS 2020 without retraining, the frozen 2015 model achieves AUC-ROC = 0.781 (drift: −0.069), ECE = 0.050 (drift: +0.039), and sensitivity = 0.267 (drift: −0.509) (Table 2). The AUC decline is moderate, indicating that the model retains reasonable discrimination; however, the sensitivity collapse at the clinical threshold is clinically catastrophic: 73% of detectable heart disease cases are missed. The frozen Platt scaler also degrades substantially (ECE 0.011 → 0.050), indicating that calibration validity is not preserved across survey years.

A full operating-point panel for the transported model at the screening threshold clarifies this failure mode. The model becomes hyper-conservative: it flags only 8.6% of patients (selection rate 0.086, against a true prevalence of 8.6%), giving high specificity (0.931) and a low false-positive rate (0.069) but recovering only 26.7% of true cases. Metrics that are insensitive to the positive class mask this collapse. Negative predictive value stays high (0.931), yet this reflects the low prevalence rather than genuine ruling-out skill, since a trivial predict-all-negative rule would reach a comparable value. Balanced accuracy is 0.599, barely above chance, and positive predictive value is 0.266, so even among flagged patients only about one in four has the outcome. Net benefit at a 0.12 threshold remains slightly positive (0.014), exceeding both the treat-all and treat-none baselines, but its small magnitude confirms that the transported model offers limited clinical value until retraining restores sensitivity.

Retraining on BRFSS 2020 restores AUC = 0.840, ECE = 0.012, and sensitivity = 0.731. The near-complete ECE recovery and partial sensitivity recovery confirm that degradation reflects five-year distribution shift rather than intrinsic difficulty of the 2020 cohort. Meta-learner coefficients shift substantially under retraining: XGBoost receives a negative coefficient (−0.427), LGBM becomes dominant (3.510), and MLP drops to third (1.454), demonstrating that the relative contribution of base learners is dataset-dependent and that full-stack retraining is required when survey characteristics change.

Race/ethnicity fairness analysis on the retrained 2020 model reveals a TPR gap of 0.206 across the six racial groups. Hispanic and Asian patients have the lowest TPRs (0.564 and 0.542 respectively), while White patients have the highest (0.748). Under temporal transport (where race is not a training feature), the race TPR gap is 0.117; because the transported model operates at sharply reduced sensitivity (0.267), this lower gap does not by itself isolate the effect of race as a feature, which we test directly through a controlled ablation below. SHAP temporal stability analysis confirms that Age and GenHealth retain their top-2 positions across both survey years, with Age importance increasing (0.764 → 0.981), making the retrained 2020 model even more age-dependent.

**Fig. 10.**
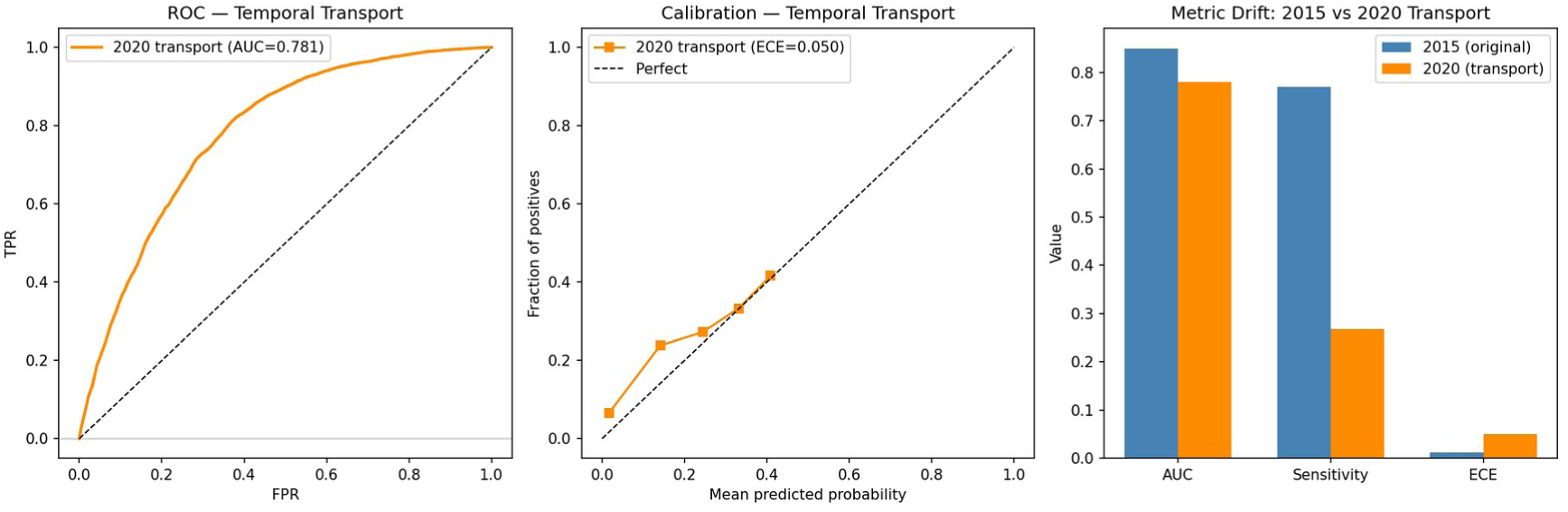
Temporal drift analysis: ROC curve on BRFSS 2020 under transport (left), calibration drift (centre), and metric comparison bar chart (right). AUC decline is moderate (−0.071) but sensitivity collapse at pt = 0.12 is clinically critical (0.776 → 0.267, annotated).

**Fig. 11.**
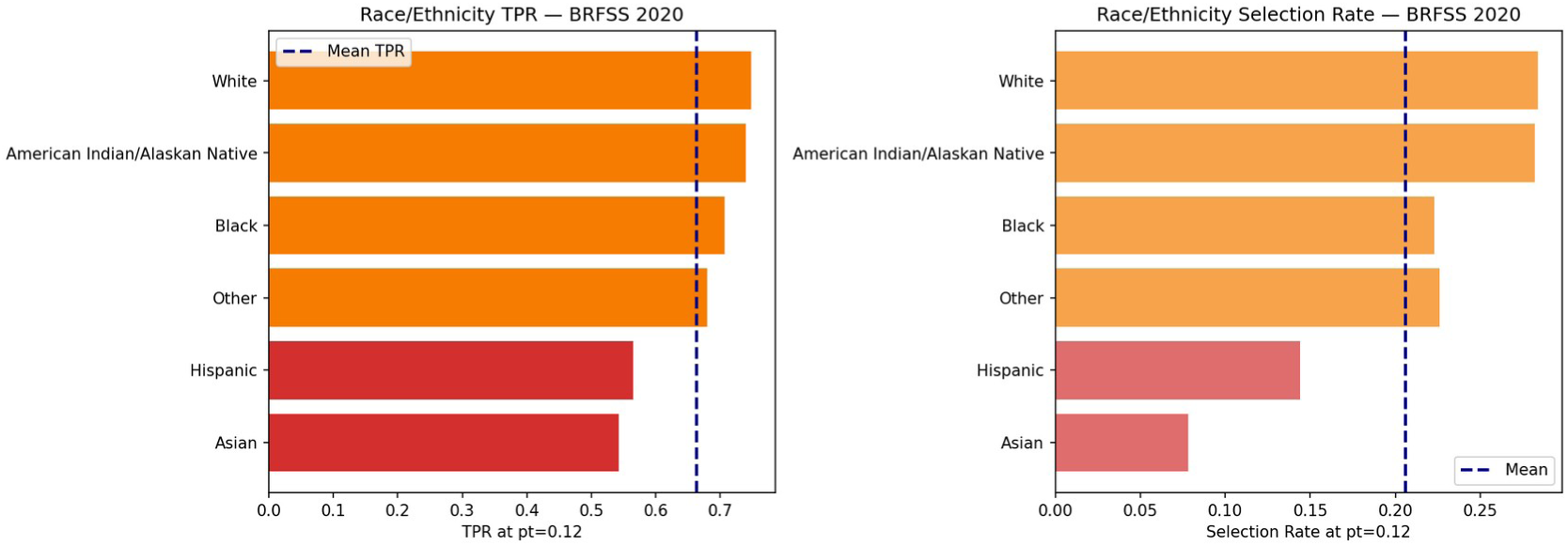
Race/Ethnicity TPR (left) and selection rate (right) on the retrained BRFSS 2020 model. Hispanic (0.564) and Asian (0.542) patients face the largest TPR deficits relative to White patients (0.748).

### 5.5. Cardio Dataset: Cross-Domain Validation

On the Cardiovascular Disease clinical examination dataset (n = 13,746 test set; 49.5% positive), the stacked ensemble achieves AUC-ROC = 0.807, AUPRC = 0.788, and Brier score = 0.180 at pt = 0.50 (Table 2). The lower AUC compared with BRFSS (0.807 vs 0.850) reflects the greater classification difficulty of clinical exam data: unlike BRFSS, where elderly multi-morbid patients dominate the positive class and are easily identified, the Cardio dataset’s age range (29.6–64.9) and balanced prevalence (49.5%) present a more uniform classification challenge.

Notably, the raw stack is already near-perfectly calibrated before Platt scaling (ECE = 0.018), and Platt scaling marginally worsens calibration (ECE = 0.027). This context-dependent behaviour contrasts sharply with BRFSS, where Platt scaling was essential. The difference is attributable to the balanced class distribution in Cardio, which removes the systematic probability underestimation that Platt scaling corrects in imbalanced settings.

SHAP analysis identifies systolic BP (ap_hi) as the dominant feature (mean |SHAP| = 0.848), followed by age (0.283) and cholesterol (0.249). SHAP–LIME consistency analysis reveals a cross-domain inversion: Kendall-τ = 0.485 (p = 0.031, significant) but Jaccard@3 = 1.000 (perfect top-3 overlap: ap_hi, age, cholesterol). This pattern, lower global rank correlation but perfect top-3 agreement, contrasts with BRFSS (τ = 0.580, Jaccard@3 = 0.200) and arises because the three dominant clinical features carry such strong, unambiguous signal that both methods agree on them completely, while the remaining near-zero features are ranked in essentially random order by both, generating noise-level global rank disagreement.

**Fig. 12.**
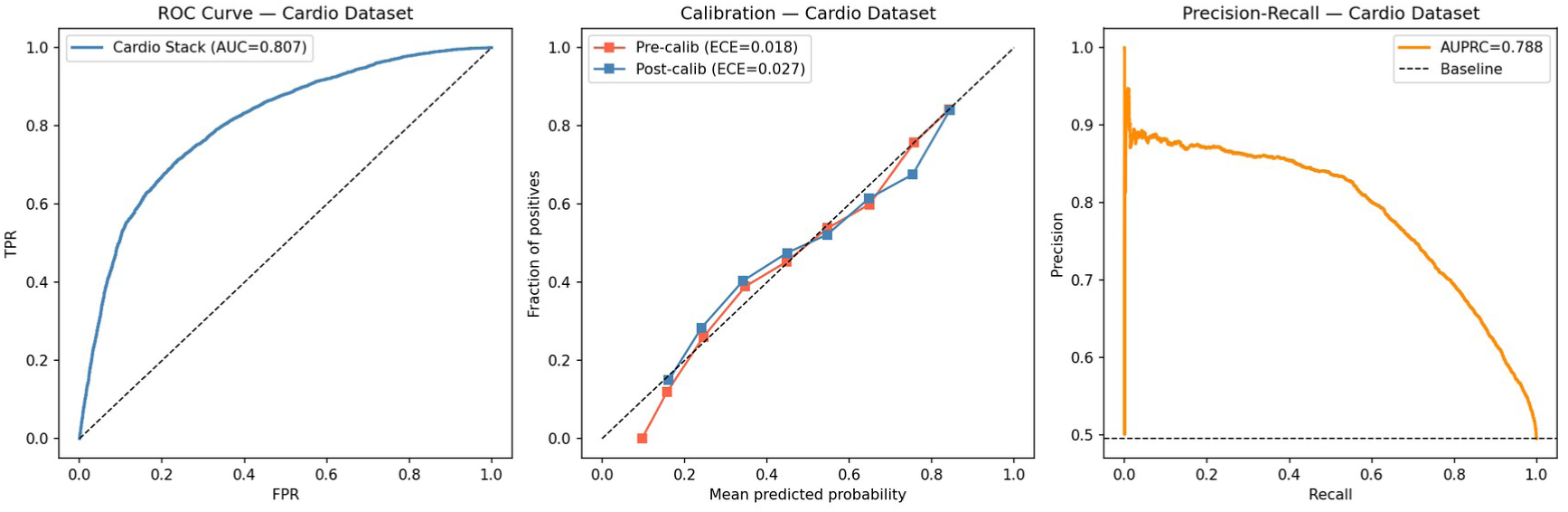
ROC curve (AUC = 0.807), Precision-Recall curve (AUPRC = 0.788), and calibration curves on the Cardiovascular Disease dataset. The raw stack is already near-perfectly calibrated (ECE = 0.018); Platt scaling marginally worsens ECE on this balanced dataset.

The gender TPR gap collapses to 0.014 on clinical examination data (Female TPR = 0.682, Male TPR = 0.696), compared with 0.124–0.164 on BRFSS. Combined with the finding that gender ranks last in SHAP importance (mean |SHAP| = 0.016 vs 0.355 in BRFSS), this indicates that female under-detection in BRFSS-based models is instrument-dependent: when objective clinical measurements replace self-reported health indicators, the sex disparity is not reproduced, consistent with a measurement or outcome-definition explanation rather than a substantive risk difference.

**Fig. 13.**
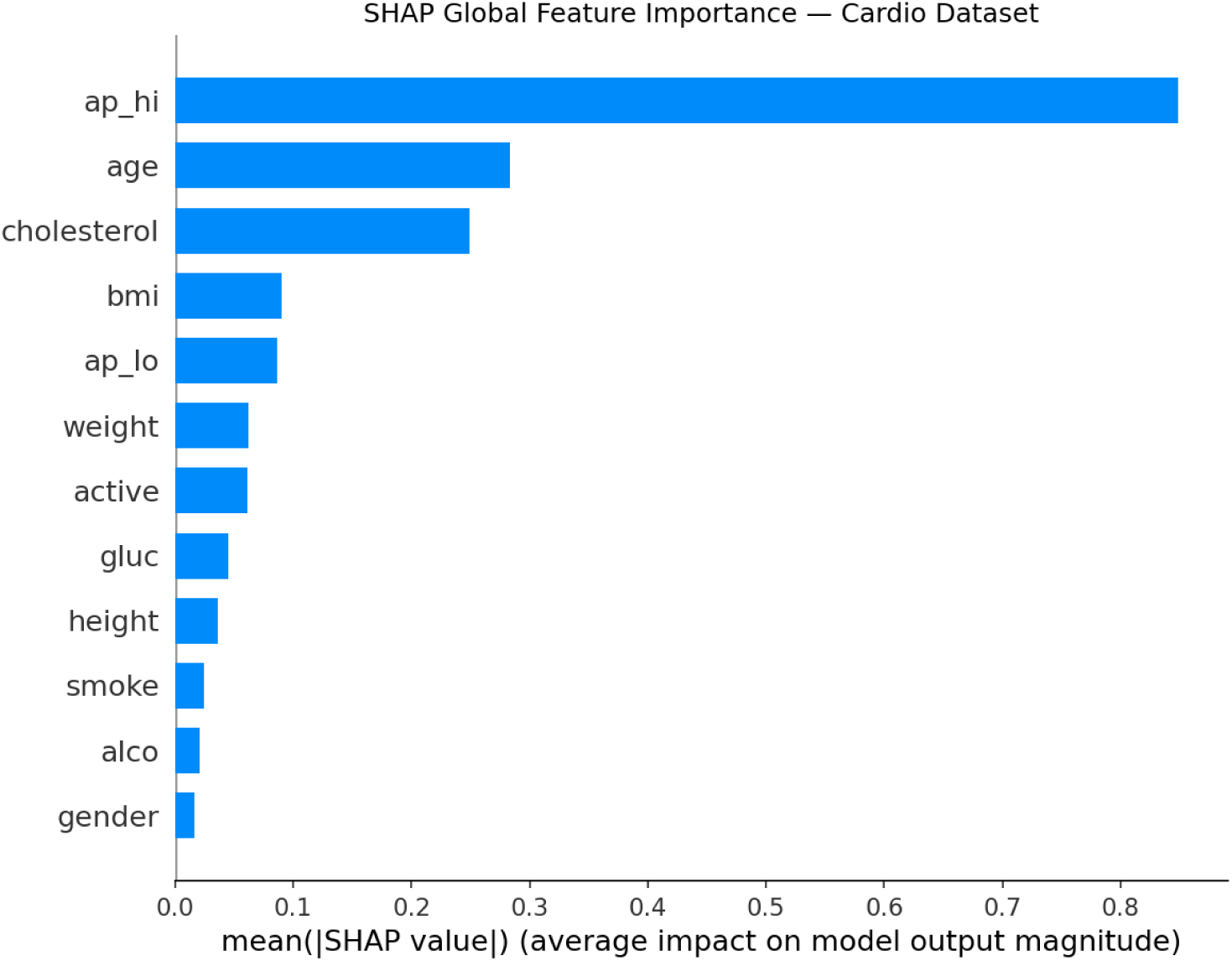
Global SHAP feature importance on the Cardiovascular Disease dataset. Systolic BP (ap_hi) dominates (mean |SHAP| = 0.848). Gender ranks last (0.016), contrasting sharply with its BRFSS rank of 4.

**Fig. 14.**
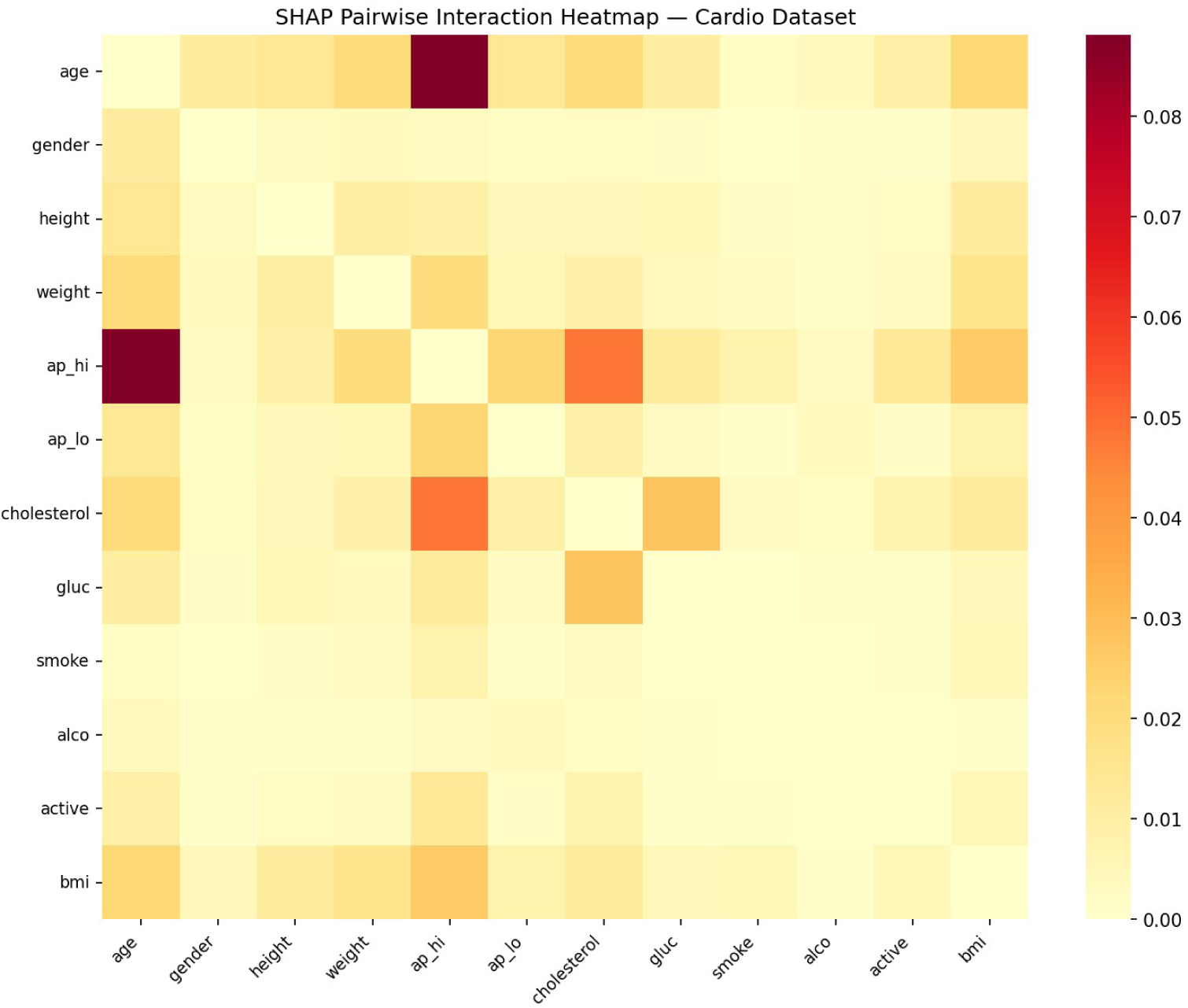
Pairwise SHAP interaction heatmap on the Cardiovascular Disease dataset. The ap_hi × ap_lo interaction is the strongest pairwise effect, followed by ap_hi × age.

**Fig. 15.**
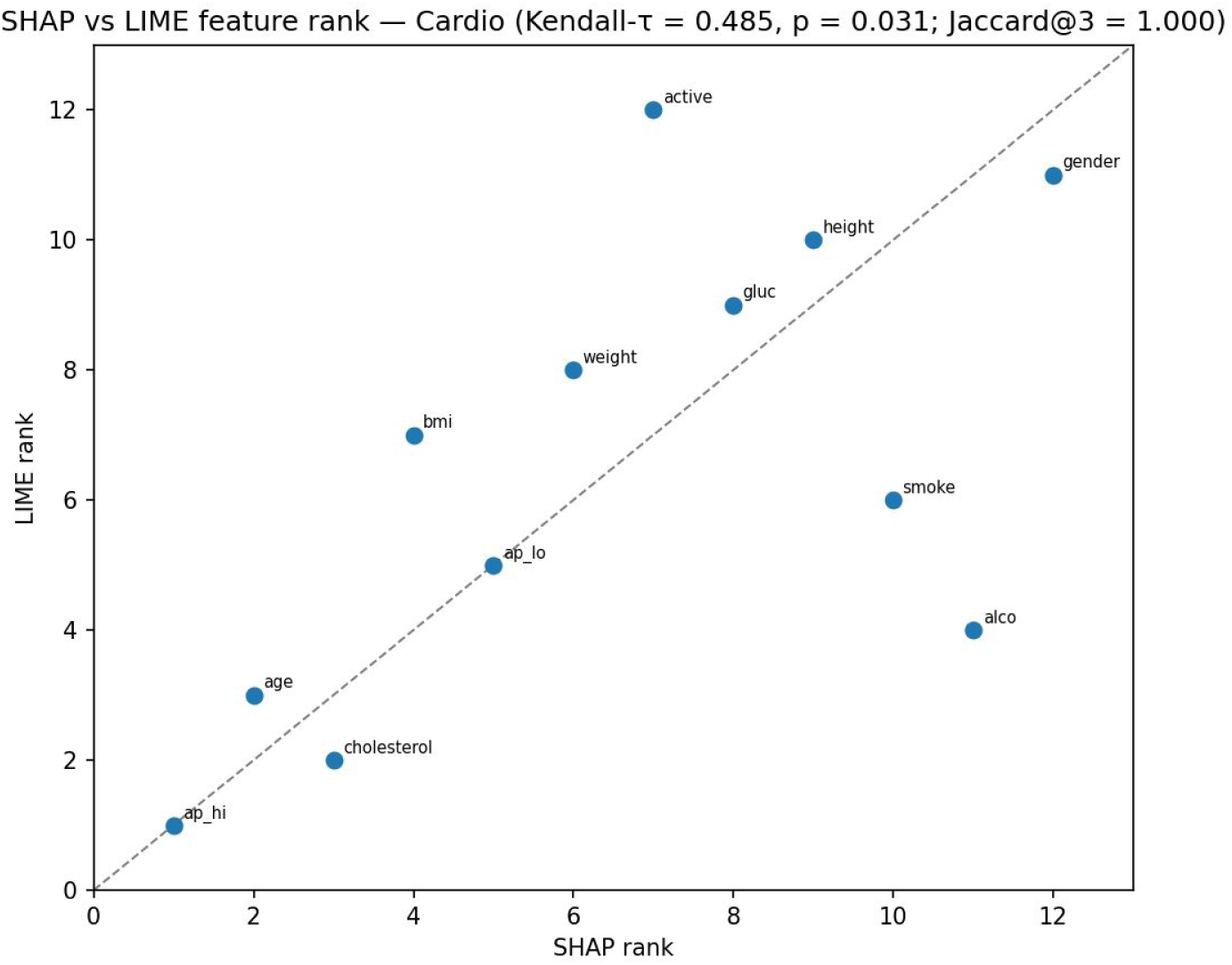
SHAP vs LIME rank scatter plot on the Cardiovascular Disease dataset (Kendall-τ = 0.485, p = 0.031; Jaccard@3 = 1.000). A cross-domain inversion of the BRFSS pattern.

**Fig. 16.**
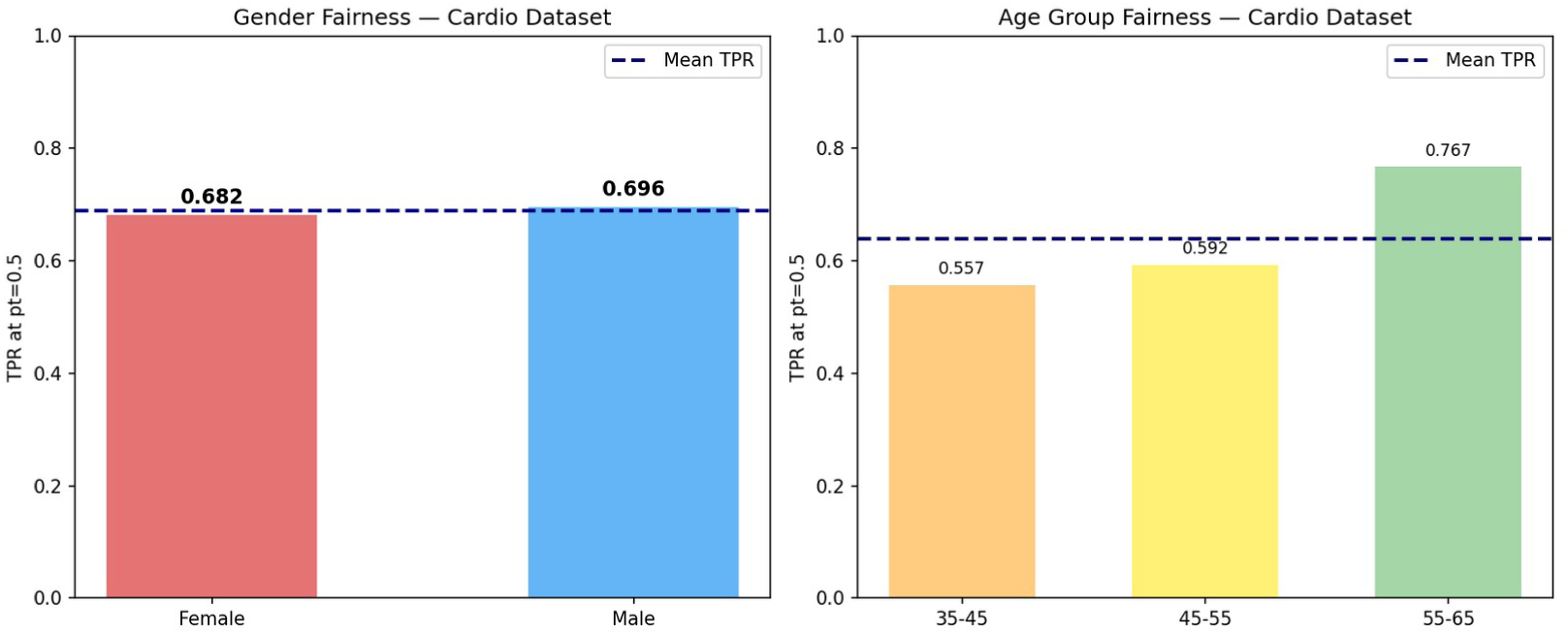
Gender fairness (left) and Age group fairness (right) on the Cardiovascular Disease dataset at pt = 0.50. Gender TPR gap = 0.014 vs 0.124–0.164 on BRFSS.

**Table 3.** SHAP–LIME consistency metrics.

| Metric | BRFSS 2015 | Cardio |
| --- | --- | --- |
| Kendall-τ | 0.580 | 0.485 |
| p-value | < 0.001 | 0.031 |
| Spearman-ρ | 0.818 | 0.622 |
| Jaccard@3 | 0.200 | 1.000 |
| Jaccard@5 | 0.429 | 0.667 |
| SHAP rank 1 | Age | ap_hi (systolic BP) |
| LIME rank 1 | Stroke | ap_hi (systolic BP) |

**Table 4.** Fairness audit results at operating threshold.

| Metric | BRFSS 2015 | 2020 Transport | 2020 Retrained | Cardio |
| --- | --- | --- | --- | --- |
| Sex/Gender TPR gap | 0.124 | 0.220 | 0.164 | 0.014 |
| Female TPR | 0.705 | 0.138 | 0.636 | 0.682 |
| Male TPR | 0.829 | 0.358 | 0.800 | 0.696 |
| Age TPR range | 0.000–0.900 | 0.000–0.467 | 0.074–0.912 | 0.557–0.767 |
| Race/Ethnicity TPR gap | N/A | 0.117 | 0.206 | N/A |
| Sex×Age max TPR gap | 0.649 | — | — | — |
| Income TPR gap | 0.230 | — | — | — |
| Post-mitigation Sex gap | 0.000 | — | — | — |

TPR = true positive rate; Sex×Age max TPR gap = maximum TPR difference across Sex×Age intersectional cells with at least 30 positive cases. — = not evaluated.
The intersectional Sex × Age gap is reported with a bootstrap 95% confidence interval (B = 1000, seed 42); per-subgroup positive counts and Wilson 95% confidence intervals for the BRFSS 2015 fairness metrics are provided in the Supplementary Material (Tables S1 and S2).

## 6. Cross-Dataset and Temporal Analysis

### 6.1. Cross-Dataset Performance Comparison

Table 2 summarises performance across all four experimental conditions. AUC-ROC is consistent across BRFSS conditions (0.781–0.850), demonstrating that the EXHEART architecture generalises within the survey instrument family. The Cardio AUC (0.807) is lower, reflecting genuine dataset difficulty rather than architectural limitation. Platt calibration is universally effective on BRFSS (ECE 0.256–0.267 → 0.011–0.012) but context-dependent on balanced clinical data (ECE 0.018 → 0.027). The temporal transport experiment provides the most clinically informative result: AUC preservation (0.850 → 0.781) coupled with sensitivity collapse (0.776 → 0.267) illustrates the gap between aggregate discrimination and threshold-dependent clinical utility in deployed models.

**Fig. 17.**
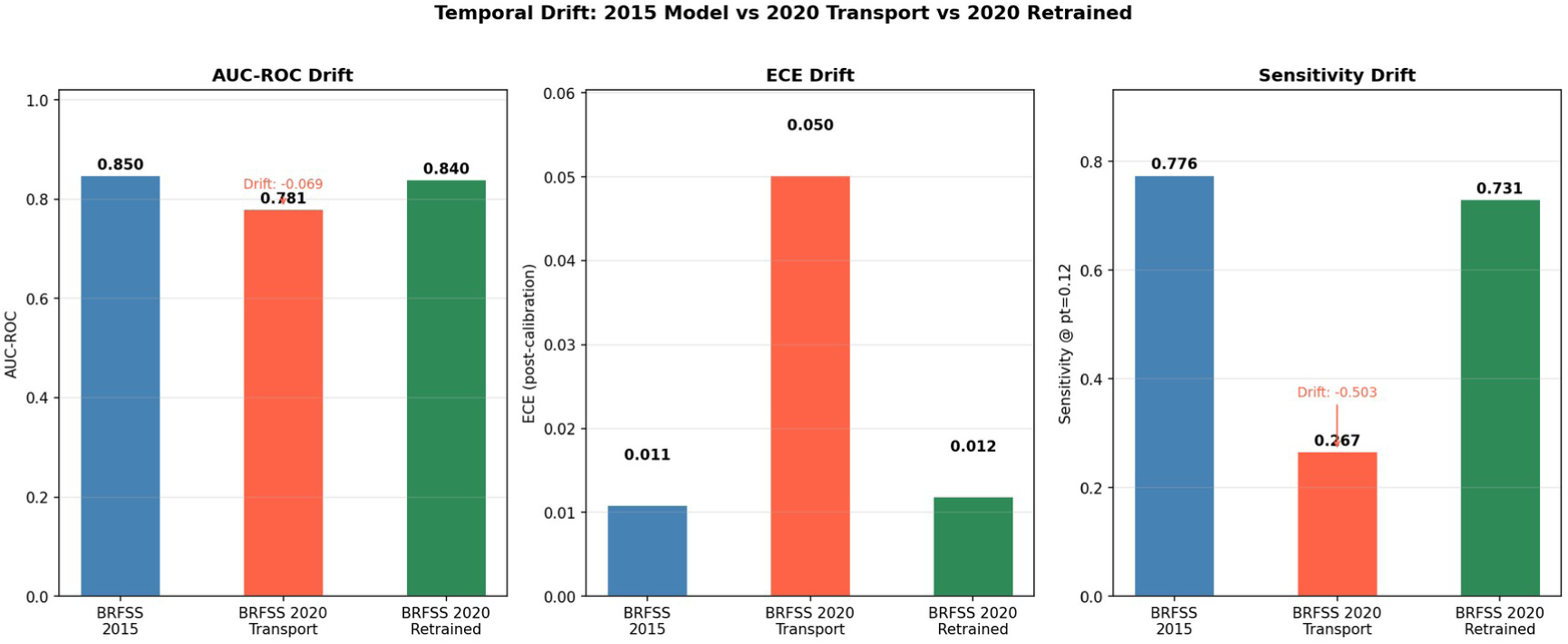
Temporal drift summary: AUC-ROC, ECE, and Sensitivity across BRFSS 2015, BRFSS 2020 transport, and BRFSS 2020 retrained conditions. The sensitivity collapse under transport (annotated, Drift: −0.509) is the critical deployment finding.

### 6.2. SHAP Temporal and Cross-Domain Stability

Comparing SHAP global rankings across BRFSS 2015 and BRFSS 2020 for the eleven matched features, Age and GenHealth are perfectly stable at ranks 1 and 2. Diabetes rises from rank 10 to rank 5 in 2020, potentially reflecting the increased prevalence of Type 2 diabetes in the U.S. adult population between survey waves. Smoker and DiffWalk show minor shifts (≤ 2 ranks), consistent with stable lifestyle-related CVD risk. Cross-domain SHAP portability analysis identifies four instrument-independent features: age (ranks 1 → 2), systolic BP/HighBP (ranks 3 → 1), cholesterol/HighChol (ranks 5 → 3), and glucose/diabetes (ranks 10 → 8). These represent the core CVD risk factor set whose relative importance is robust to measurement modality. Two major portability failures are identified: BMI rises from rank 12 to rank 4 (measured BMI substantially more informative than self-reported coded BMI), and Sex/gender falls from rank 4 to rank 12 (gender adds negligible independent information when objective clinical measurements are available).

**Fig. 18.**
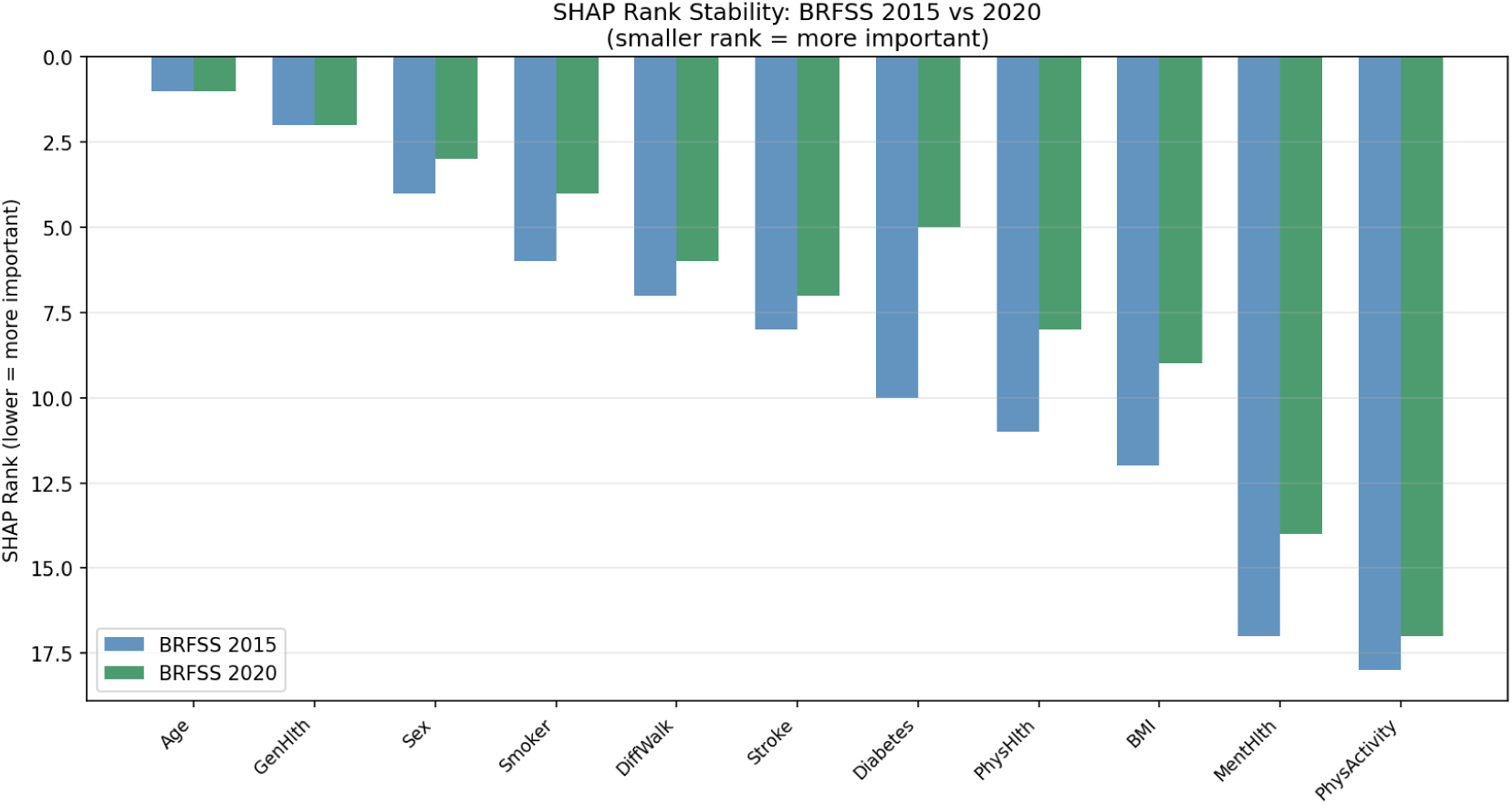
SHAP rank stability: BRFSS 2015 (blue) vs BRFSS 2020 retrained (green) for 11 matched features. Age and GenHlth are perfectly stable at ranks 1–2. Diabetes rises notably (+5 ranks) in 2020.

**Fig. 19.**
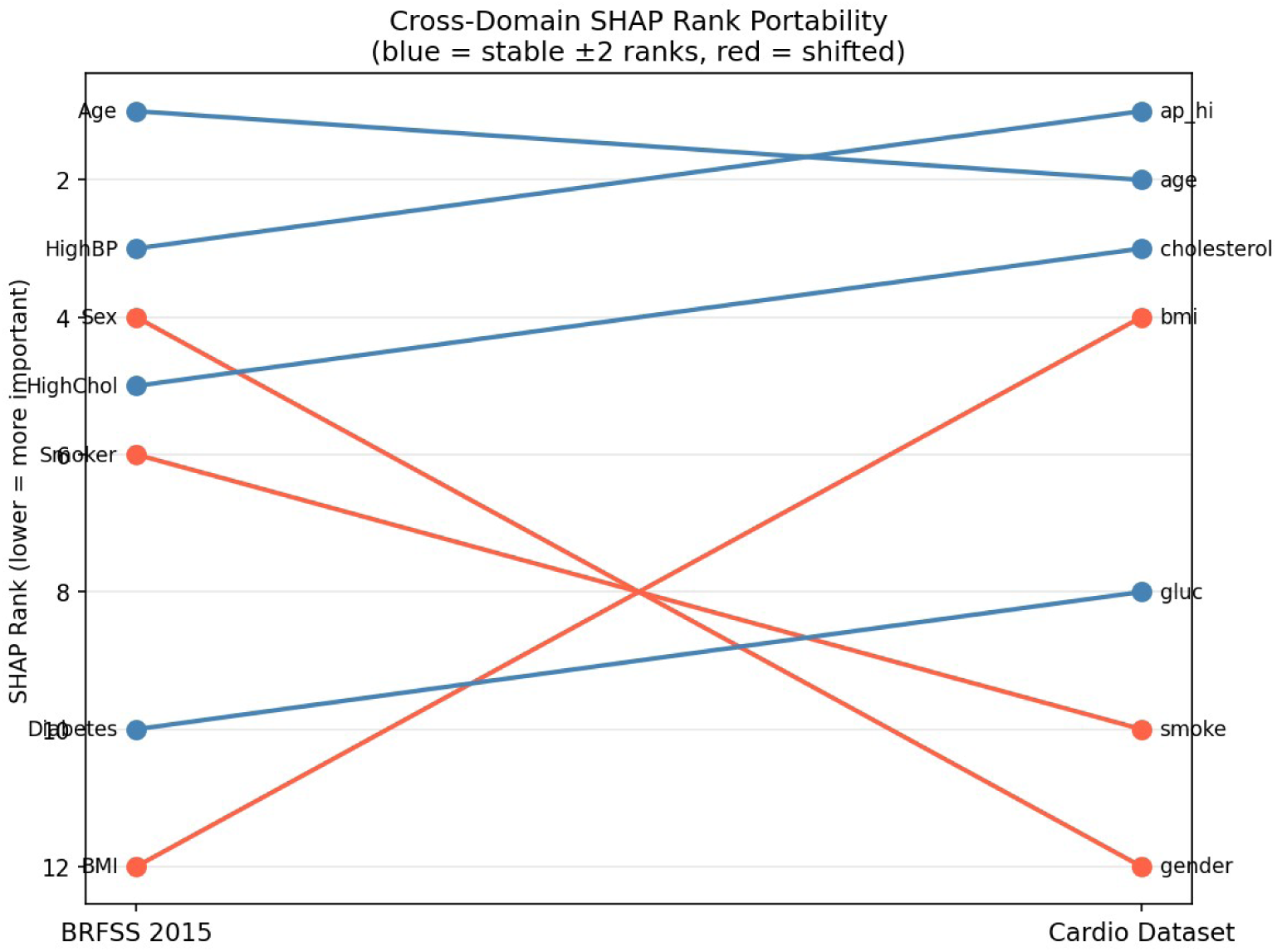
Cross-domain SHAP rank portability slope graph (BRFSS 2015 → Cardiovascular Disease). Blue = stable features (±2 ranks): Age, ap_hi/HighBP, cholesterol/HighChol, glucose/Diabetes. Red = major portability failures: BMI rises 8 ranks (measured vs self-reported), Sex/gender falls 8 ranks (clinically redundant when objective measurements available).

### 6.3. Cross-Dataset Fairness Comparison

Three cross-dataset fairness narratives emerge from the analysis. First, the Sex/gender TPR gap evolves as: 0.124 (BRFSS 2015) → 0.220 (BRFSS 2020 transport) → 0.164 (BRFSS 2020 retrained) → 0.014 (Cardio). The gap widens under temporal transport without retraining (confirming that deployed models amplify existing disparities), partially recovers with retraining, and near-completely disappears on clinical examination data. This trajectory indicates female under-detection is instrument-dependent, consistent with a measurement or outcome-definition explanation, with direct implications for the clinical deployment of BRFSS-trained screening models. Second, the age TPR gap is universal but severity varies (0.900 on BRFSS 2015 vs 0.210 on Cardio), with the difference primarily attributable to the Cardio dataset’s exclusion of patients under 30 or over 65 rather than to clinical data being intrinsically fairer. Third, the effect of the race feature is isolated through a controlled ablation: retraining the identical 2020 pipeline with versus without race as an input, holding the data, split, seeds, calibration, and threshold fixed, raises the race TPR gap from 0.143 to 0.195, a difference of 0.052 (95% CI [0.001, 0.140]). A one-hot robustness check, encoding race as indicator variables rather than an ordinal code, gives a consistent direction but a smaller, non-significant difference (0.140 without race versus 0.184 with race, difference 0.045, 95% CI [−0.009, 0.116]). The point estimate is therefore consistent across encodings, but its statistical significance depends on the encoding scheme; we accordingly interpret this as evidence that including race does not resolve, and is associated with a modest widening of, the disparity rather than as a robustly significant amplification. This same-data, single-feature comparison holds the training year and operating point fixed, unlike a transport-versus-retrain contrast in which the transported model’s collapsed sensitivity (0.267) makes its lower 0.117 gap non-comparable; the canonical retrained race gap is 0.206.

**Fig. 20.**
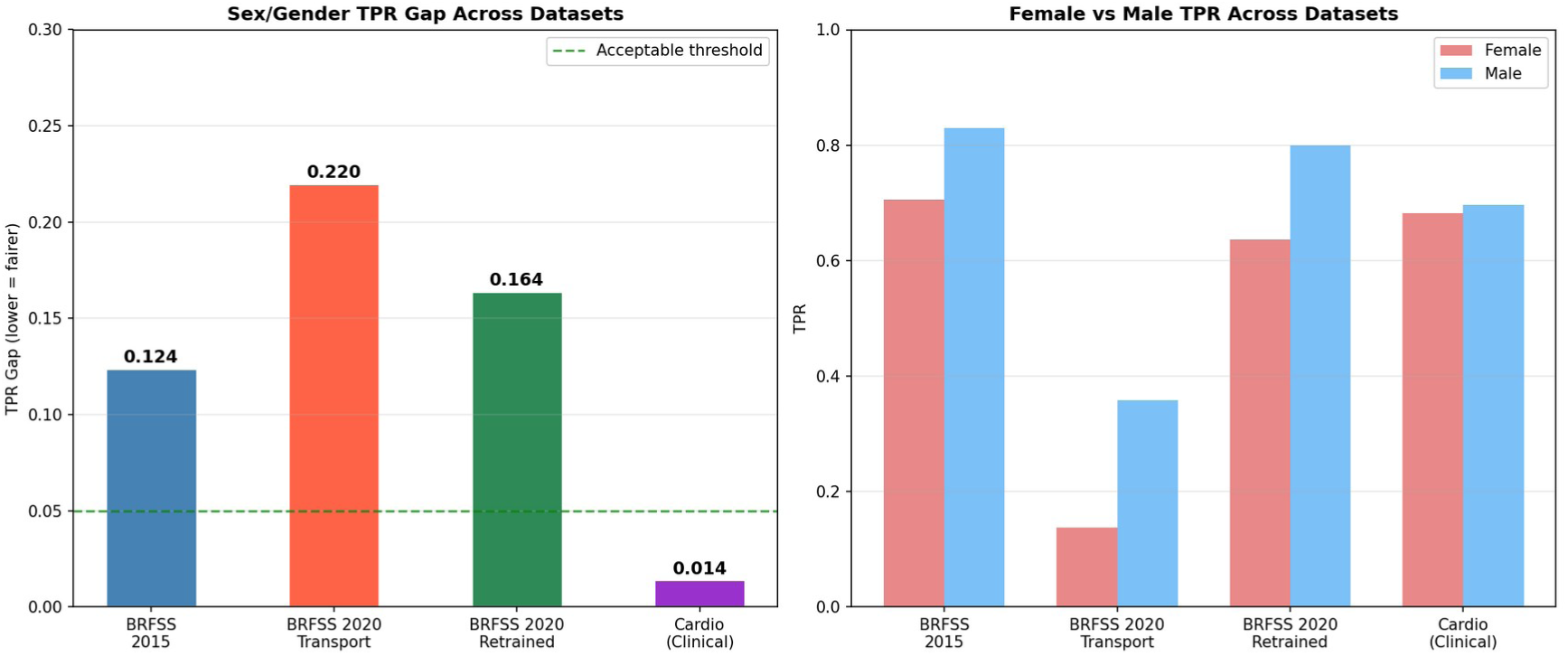
Cross-dataset Sex/Gender TPR gap (left) and Female vs Male TPR (right) across all experimental conditions. Gap trajectory: 0.124 → 0.220 → 0.164 → 0.014. Near-equality on clinical data indicates female under-detection is instrument-dependent, consistent with a measurement or outcome-definition explanation.

## 7. Discussion

### 7.1. Clinical Implications

The principal clinical implication of the temporal transport finding is that deployed BRFSS-based screening tools warrant periodic monitoring, recalibration, and retraining rather than static deployment. A model whose sensitivity collapses from 0.776 to 0.267 under combined temporal drift and feature mismatch is clinically harmful: it would miss three in four heart disease cases it would previously have detected, while preserving moderately acceptable AUC statistics that could obscure the problem from routine monitoring. We therefore recommend an explicit retraining cadence (for example, annual retraining aligned with the BRFSS survey cycle) and a drift alarm that triggers re-evaluation when sensitivity falls below a clinically defined floor.

The sex fairness attribution finding has immediate implications for model deployment in clinical decision support. If female under-detection reflects self-reporting differences in symptom perception or health-status rating rather than true population-level risk differences, then deploying BRFSS models in settings where the output is used to prioritise clinical referrals will systematically disadvantage female patients. Institutions deploying such models should either apply group-specific thresholds (which our frontier analysis shows can near-eliminate the Sex TPR gap with balanced accuracy and AUC essentially unchanged, at the cost of a modest sensitivity-for-specificity rebalancing) or validate the model against a clinical dataset before deployment.

Including race as a feature does not resolve the disparity and, under the primary (label) encoding, is associated with a modest, encoding-sensitive widening of it (controlled ablation: race TPR gap 0.143 versus 0.195 under label encoding, difference 0.052, 95% CI [0.001, 0.140]; 0.140 versus 0.184 under one-hot encoding, difference 0.045, 95% CI [−0.009, 0.116]). This argues against incorporating race as a training feature without simultaneous fairness-constrained training.

### 7.2. Methodological Contributions

The quantitative SHAP–LIME consistency framework introduced in this paper addresses a gap in clinical explainability evaluation that has persisted despite the widespread adoption of both methods. The Stroke rank inversion (rank 8 in SHAP, rank 1 in LIME) on BRFSS 2015 illustrates a scenario where a clinician using SHAP-based explanations would deprioritise stroke history as a factor in a patient’s risk assessment, while a LIME-based explanation would highlight it as the primary driver. Quantifying this divergence, and identifying the patient subgroups and features where it is largest, is a prerequisite for responsible clinical deployment of post-hoc explainability tools. The cross-domain inversion (lower τ but perfect Jaccard@3 on clinical data) demonstrates that the appropriate evaluation framework depends on the data modality: on high-signal clinical data, top-feature agreement is the relevant metric; on survey data with distributed feature importance, global rank correlation is more informative.

**Fig. 21.**
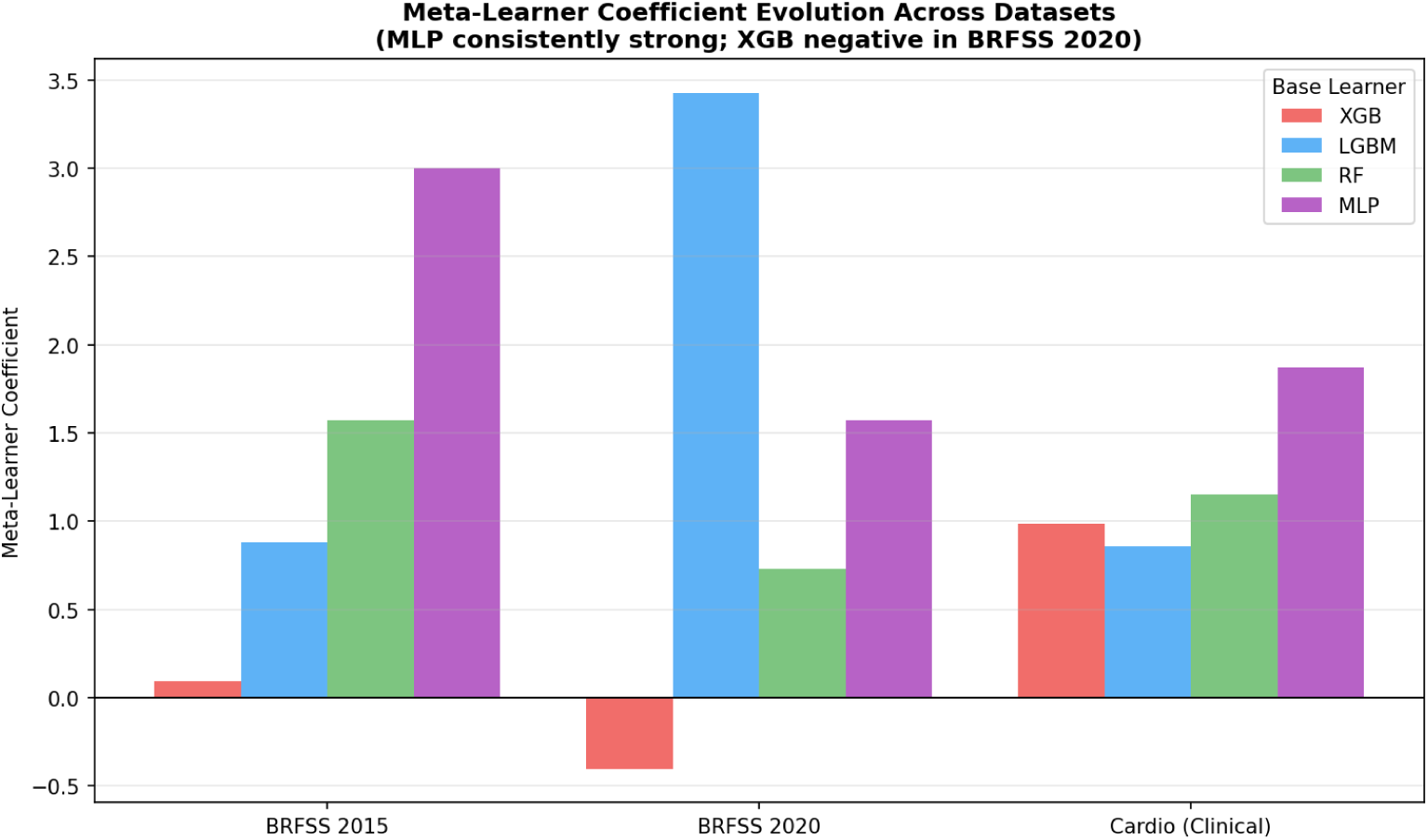
Meta-learner coefficient evolution across datasets. MLP (purple) consistently receives the highest or second-highest weight. XGBoost (red) receives a negative coefficient in BRFSS 2020, indicating its predictions are redundant given the other three models on that dataset.

The leakage-free stacking methodology delivers a 96% ECE reduction through Platt scaling while maintaining AUC = 0.850, demonstrating that anti-leakage design and strong calibration are complementary rather than competing objectives. The meta-learner coefficient analysis provides an interpretable account of base learner contributions across datasets, showing that XGBoost’s negative coefficient in BRFSS 2020 (−0.427) reflects genuine redundancy given the other models on that dataset, a signal that survives only if the full stacking architecture is evaluated, not if any single base learner is selected.

**Fig. 22.**
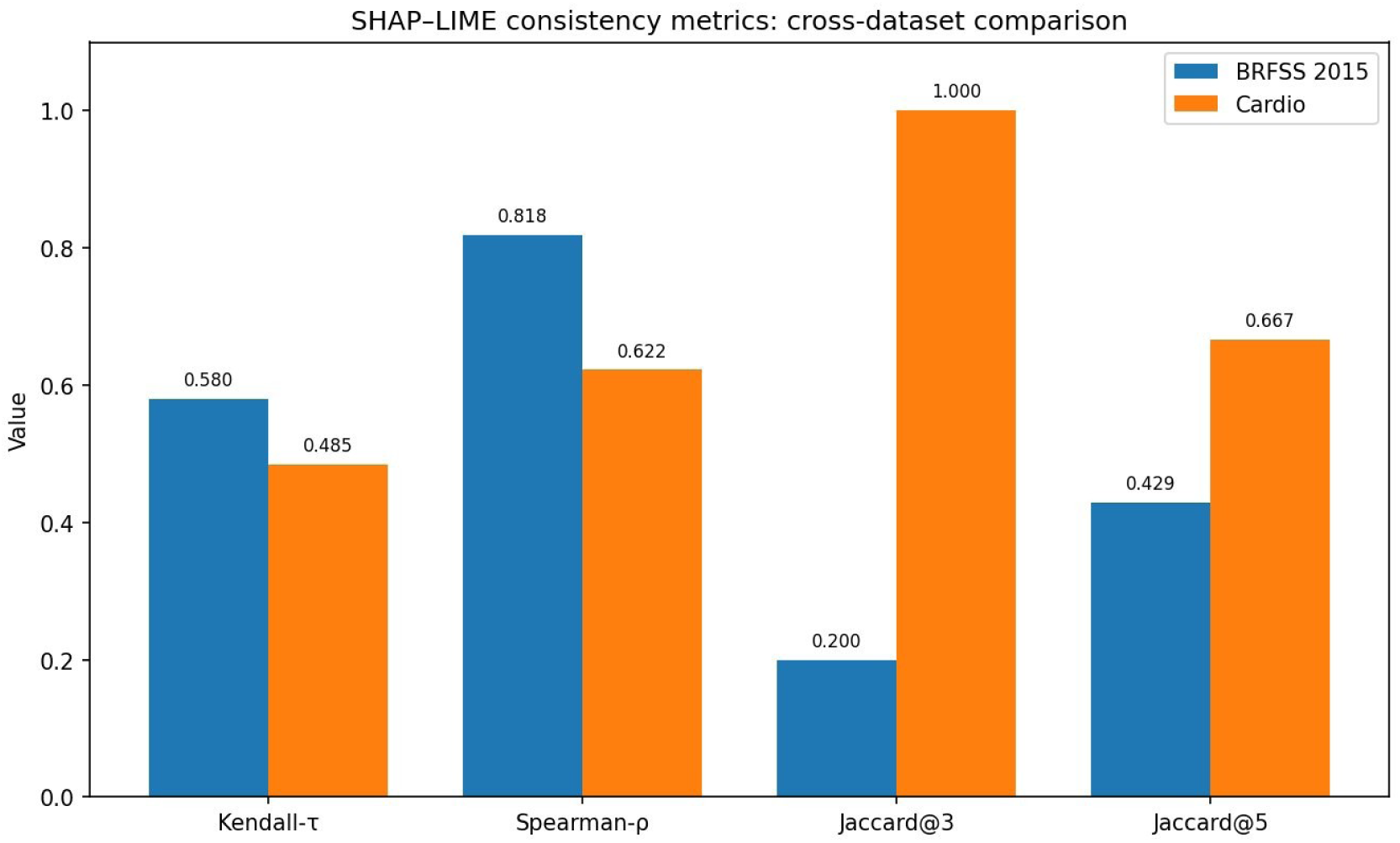
SHAP–LIME consistency metrics cross-dataset comparison. Global rank correlation is comparable across datasets (Kendall-τ = 0.580 on survey data, 0.485 on clinical data), while top-3 overlap inverts sharply, rising from 0.200 on survey data to 1.000 on clinical data. The contrast reflects high signal-to-noise ratio in clinical features vs distributed importance in survey features.

### 7.3. Distinguishing Measurement-Induced from Substantive Disparities

A recurring critique of algorithmic fairness in clinical machine learning is that disparity metrics quantify the size of a gap but reveal nothing about its cause, leaving practitioners without the grounding needed to choose an appropriate intervention; recent work argues that researchers should step outside the fairness-metric frame and engage with the mechanisms that actually produce health disparities [32], a position echoed in calls for explicit causal reasoning about their sources [33]. The cross-instrument attribution test introduced in this paper responds to that critique. By re-evaluating a survey-derived disparity on a dataset that measures the same clinical construct objectively, the test separates disparities that arise from how the data are collected from those that reflect genuine differences in risk between groups.

The female under-detection disparity illustrates the diagnostic. On both survey waves the Sex TPR gap is large (0.124 in 2015 and 0.164 in the retrained 2020 model) and Sex is the fourth most important SHAP feature. On the clinical examination dataset the same gap falls to 0.014 and Sex drops to the least important of twelve features. The simultaneous collapse of the disparity and of the responsible attribution is the signature of a measurement-induced gap: the predictive weight the model places on sex when trained on survey data appears to stand in for information that objective blood-pressure, cholesterol, and BMI measurements supply directly once they are available. The remedy is concrete: rather than constrain the model, recognise that a survey-trained screening tool will systematically under-detect women, and either deploy it with sex-specific thresholds or validate it against objectively measured data before use.

Three controlled checks corroborate this attribution. First, restricting both datasets to the overlapping 30 to 64 age band leaves the survey sex gap essentially unchanged (0.112 versus 0.124 for all ages), which rules out an age-cohort confound. Second, a bootstrap analysis (B = 1000 resamples) estimates that 89% of the survey sex gap is removed on clinical measurement, with a 95% confidence interval of [0.65, 0.99] that excludes zero, so the collapse is unlikely to be a sampling artefact. Third, coarsening the clinical features to survey-style binary indicators lowers discrimination (AUC 0.806 to 0.778) but does not reintroduce the sex gap (0.013 versus 0.012; difference 95% CI [−0.013, 0.015], straddling zero), which is consistent with attributing the disparity to the self-reported outcome rather than to feature granularity.

Age behaves differently, and the contrast shows why the test must be read with care. The age disparity persists across every dataset and narrows on the clinical data only because that cohort excludes the under-30 and over-65 groups that drive the survey gap, not because objective measurement resolves it. That disparity is substantive rather than an artefact, and algorithmic or threshold intervention must address it directly. The test generalises to any setting where a construct can be captured by both a subjective and an objective instrument, but its validity rests entirely on that construct match; where no objectively measured counterpart exists, attribution cannot be performed and the disparity should be treated as substantive by default. Cross-instrument attribution is therefore a complement to conventional mitigation, not a replacement: it tells the practitioner which gaps to fix in the model and which to fix upstream in data collection. We frame it as an empirical instantiation of fairness-gap attribution rather than a new decomposition method (Section 2.8); where the closest prior analog isolates an input-acquisition artefact in medical imaging, EXHEART addresses the complementary case of an outcome-labelling channel in survey-based prediction [37].

### 7.4. Limitations

Several limitations constrain the generalisability of these findings. First, BRFSS is a cross-sectional telephone survey subject to self-reporting bias, recall bias, and non-response bias; the observed fairness disparities may therefore reflect measurement inequalities as much as true population-level risk differences. Second, the Cardiovascular Disease dataset lacks patients under 30 or over 65 and contains no race/ethnicity variable, limiting the scope of the cross-domain fairness analysis. Third, the ten features imputed with 2015 medians in the temporal transport experiment (including HighBP and HighChol) are likely informative, meaning the transport degradation represents a lower bound on true deployment drift for a model deployed in an environment where those features remain unavailable. Fourth, the threshold-shifting mitigation applied in this paper is a post-hoc correction that does not alter the underlying model; more sophisticated fairness-constrained training approaches may achieve better Pareto efficiency. Fifth, the SHAP–LIME consistency analysis uses a random sample of 200 instances for LIME, which may not fully capture the distribution of explanation disagreement across the test set. Future work should examine per-stratum consistency on larger samples and explore whether the Explanation Reliability Index (an aggregated per-patient consistency score combining SHAP stability, LIME re-run Jaccard, and SHAP–LIME agreement) provides clinically actionable reliability estimates. EXHEART is intended for retrospective model evaluation and population-level cardiovascular screening research. It has not been validated for direct clinical decision-making and should not be used to guide individual diagnosis or treatment without prospective clinical validation.

## 8. Conclusion

This paper presented EXHEART, a fairness-aware, explainable stacked ensemble pipeline for cardiovascular disease classification, validated across three datasets spanning two survey waves and one clinical examination cohort. Five contributions were demonstrated. A leakage-free four-model stacking architecture with Platt calibration achieves AUC = 0.850 and reduces ECE by 96% on BRFSS 2015, with the MLP base learner receiving the highest meta-learner weight and contributing the most unique signal to the ensemble. A quantitative SHAP–LIME consistency framework, the first applied to BRFSS-scale data, reveals a clinically meaningful top-3 divergence on survey data (Jaccard@3 = 0.200) and a cross-domain inversion on clinical data (τ = 0.485, p = 0.031; Jaccard@3 = 1.000). Subgroup-stratified SHAP interaction analysis identifies differential feature interaction patterns across demographic groups. An intersectional fairness audit reveals a Sex × Age TPR gap of 0.649 among adequately-powered cells, 5.2× the single-attribute Sex gap, with a smooth fairness–utility frontier along which per-group threshold shifting can near-eliminate the Sex TPR gap (0.124 to approximately 0.000) with balanced accuracy and AUC essentially unchanged. A feature-mismatch temporal transport stress test demonstrates that sensitivity collapses from 0.776 to 0.267 without retraining, with near-complete recovery on retraining, establishing that periodic monitoring, recalibration, and retraining should be evaluated before deployment. Cross-domain analysis indicates that female under-detection is instrument-dependent and is not reproduced on clinical examination data, consistent with a measurement or outcome-definition explanation, with direct implications for the equitable deployment of population survey-based screening tools.

Future work will explore an Explanation Reliability Index that aggregates per-patient SHAP stability, LIME re-run consistency, and SHAP–LIME agreement into a single clinically actionable score, concept-level explanation portability across measurement modalities, and prospective validation of the EXHEART pipeline against longitudinal incident CVD outcomes in electronic health record cohorts.

## Data Availability

All datasets analysed in this study are openly available in public repositories. The BRFSS 2015 Heart Disease Health Indicators dataset and the Cardiovascular Disease dataset are hosted on Kaggle; the cleaned BRFSS 2020 dataset is archived on Zenodo (https://doi.org/10.5281/zenodo.15364962). The code supporting this study is available at https://github.com/anasbiswas1/exheart-research. No data were generated that are not publicly available.

https://www.kaggle.com/datasets/sulianova/cardiovascular-disease-dataset

https://github.com/anasbiswas1/exheart-research

## Ethics statement

This study analysed three publicly available, fully de-identified secondary datasets: the BRFSS 2015 and BRFSS 2020 surveys collected by the U.S. Centers for Disease Control and Prevention, and the Cardiovascular Disease examination dataset distributed via Kaggle. No identifiable personal information was accessed at any stage. Because the work relied exclusively on existing, anonymised, publicly released data and involved no contact with human participants, it did not require institutional ethical review or informed consent.

## CRediT authorship contribution statement

Md Anas Biswas: Conceptualization, Methodology, Software, Formal analysis, Investigation, Data curation, Writing – original draft, Visualization. Alif Laila: Conceptualization, Validation, Writing – review & editing.

## Data and code availability

All datasets analysed in this study are publicly available. The BRFSS 2015 Heart Disease Health Indicators subset is hosted on Kaggle [27]; the cleaned BRFSS 2020 dataset is archived on Zenodo (DOI: 10.5281/zenodo.15364962) [28]; and the Cardiovascular Disease examination dataset is hosted on Kaggle [29]. The pipeline was implemented in Python using scikit-learn, XGBoost, LightGBM, TensorFlow/Keras, the SHAP library, and LIME. The full analysis code, fixed random seeds (seed = 42), library environment specification, and notebook execution order required to reproduce every reported result are available at https://github.com/anasbiswas1/exheart-research.

## Declaration of generative AI and AI-assisted technologies in the writing process

During the preparation of this manuscript, the authors used Claude (Anthropic) to assist with drafting and language editing of the text and with the formatting of tables and figures. After using this tool, the authors reviewed and edited the content as needed, verified all reported results, and take full responsibility for the content of the publication.

## Supplementary Material

This supplement reports per-subgroup positive counts and 95% Wilson confidence intervals for the BRFSS 2015 fairness metrics summarised in Table 4. The intersectional Sex × Age maximum gap reported in the main text (0.649, 95% CI [0.545, 0.814]) is computed over cells with at least 30 positive cases; its confidence interval is obtained by bootstrap resampling (B = 1000, seed 42). The true positive rate is undefined or unstable for cells with very few positive cases, which is why a positive-count floor is applied to the headline intersectional gap.

**Table S1.** Per-subgroup true positive rate, positive counts, and 95% Wilson confidence intervals on BRFSS 2015 at the screening threshold (single-attribute subgroups; age band 1 = youngest, 13 = oldest).

| Subgroup | n | Positives | TPR | 95% Wilson CI |
| --- | --- | --- | --- | --- |
| **Sex** |  |  |  |  |
| Female | 28300 | 2051 | 0.705 | [0.685, 0.724] |
| Male | 22436 | 2728 | 0.829 | [0.815, 0.843] |
| **Age band** |  |  |  |  |
| 1 | 1123 | 7 | 0.143 | [0.026, 0.513] |
| 2 | 1567 | 13 | 0.000 | [0.000, 0.228] |
| 3 | 2228 | 23 | 0.261 | [0.125, 0.465] |
| 4 | 2811 | 39 | 0.128 | [0.056, 0.267] |
| 5 | 3214 | 63 | 0.317 | [0.216, 0.440] |
| 6 | 4024 | 155 | 0.439 | [0.363, 0.517] |
| 7 | 5238 | 306 | 0.546 | [0.490, 0.601] |
| 8 | 6262 | 453 | 0.684 | [0.640, 0.725] |
| 9 | 6606 | 659 | 0.744 | [0.709, 0.775] |
| 10 | 6399 | 853 | 0.809 | [0.781, 0.834] |
| 11 | 4650 | 760 | 0.867 | [0.841, 0.889] |
| 12 | 3156 | 608 | 0.882 | [0.853, 0.905] |
| 13 | 3458 | 840 | 0.900 | [0.878, 0.919] |

**Table S2.**
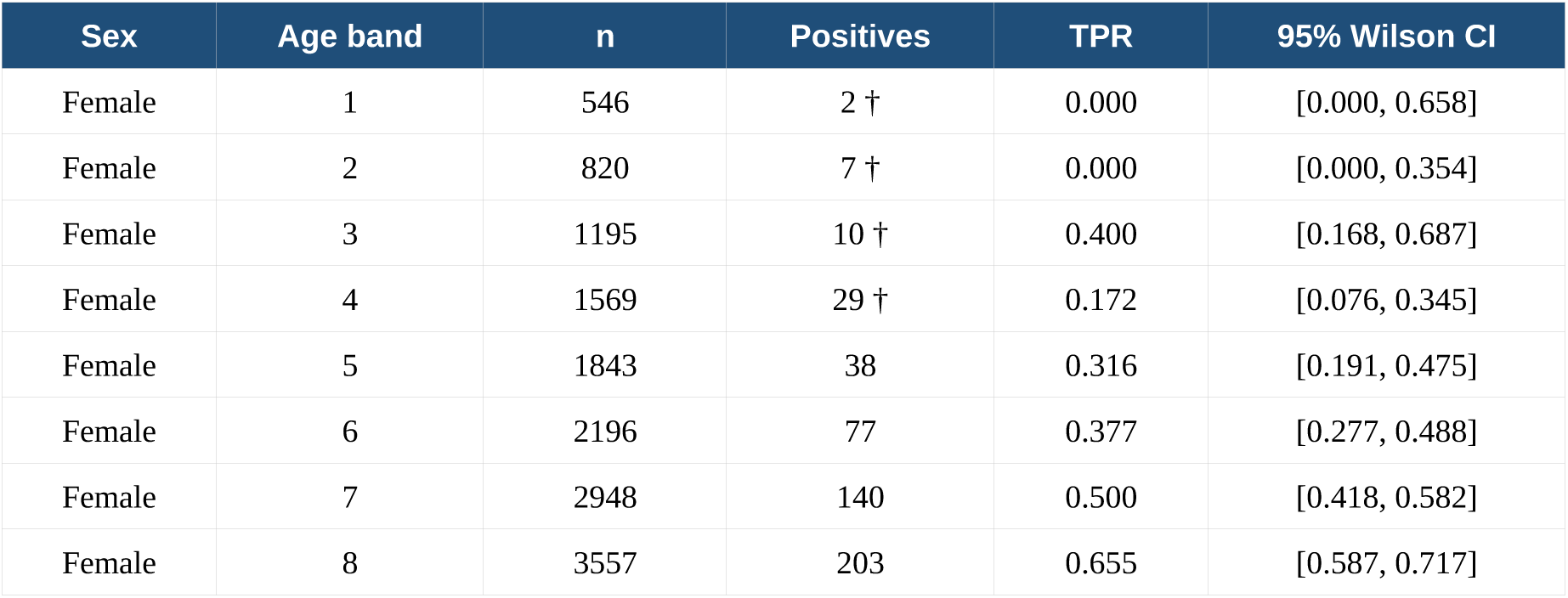
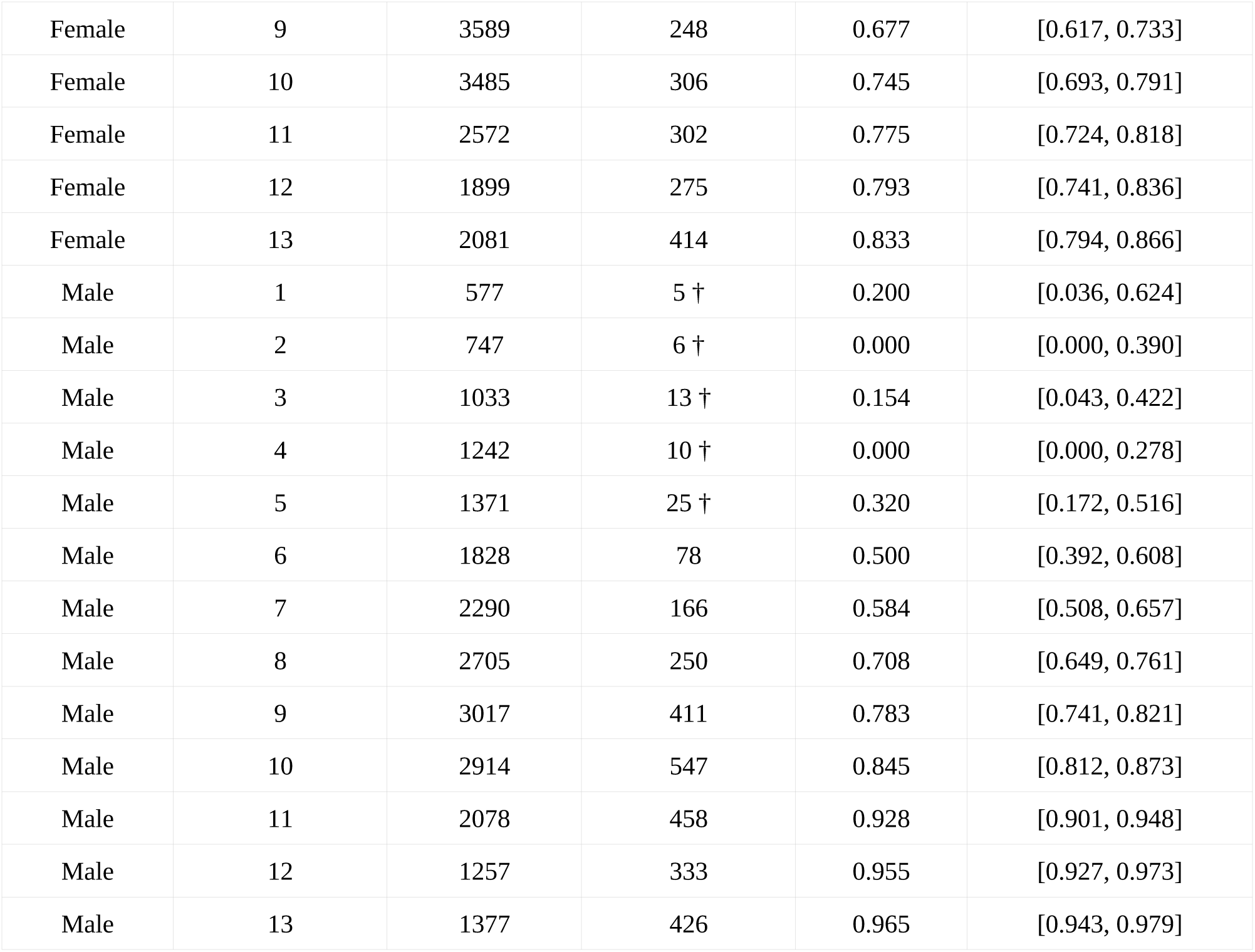
Sex × Age intersectional true positive rate, positive counts, and 95% Wilson confidence intervals on BRFSS 2015 at the screening threshold. Cells marked † contain fewer than 30 positive cases and are excluded from the headline intersectional gap.

| Sex | Age band | n | Positives | TPR | 95% Wilson CI |
| --- | --- | --- | --- | --- | --- |
| Female | 1 | 546 | 2 † | 0.000 | [0.000, 0.658] |
| Female | 2 | 820 | 7 † | 0.000 | [0.000, 0.354] |
| Female | 3 | 1195 | 10 † | 0.400 | [0.168, 0.687] |
| Female | 4 | 1569 | 29 † | 0.172 | [0.076, 0.345] |
| Female | 5 | 1843 | 38 | 0.316 | [0.191, 0.475] |
| Female | 6 | 2196 | 77 | 0.377 | [0.277, 0.488] |
| Female | 7 | 2948 | 140 | 0.500 | [0.418, 0.582] |
| Female | 8 | 3557 | 203 | 0.655 | [0.587, 0.717] |
| Female | 9 | 3589 | 248 | 0.677 | [0.617, 0.733] |
| Female | 10 | 3485 | 306 | 0.745 | [0.693, 0.791] |
| Female | 11 | 2572 | 302 | 0.775 | [0.724, 0.818] |
| Female | 12 | 1899 | 275 | 0.793 | [0.741, 0.836] |
| Female | 13 | 2081 | 414 | 0.833 | [0.794, 0.866] |
| Male | 1 | 577 | 5 † | 0.200 | [0.036, 0.624] |
| Male | 2 | 747 | 6 † | 0.000 | [0.000, 0.390] |
| Male | 3 | 1033 | 13 † | 0.154 | [0.043, 0.422] |
| Male | 4 | 1242 | 10 † | 0.000 | [0.000, 0.278] |
| Male | 5 | 1371 | 25 † | 0.320 | [0.172, 0.516] |
| Male | 6 | 1828 | 78 | 0.500 | [0.392, 0.608] |
| Male | 7 | 2290 | 166 | 0.584 | [0.508, 0.657] |
| Male | 8 | 2705 | 250 | 0.708 | [0.649, 0.761] |
| Male | 9 | 3017 | 411 | 0.783 | [0.741, 0.821] |
| Male | 10 | 2914 | 547 | 0.845 | [0.812, 0.873] |
| Male | 11 | 2078 | 458 | 0.928 | [0.901, 0.948] |
| Male | 12 | 1257 | 333 | 0.955 | [0.927, 0.973] |
| Male | 13 | 1377 | 426 | 0.965 | [0.943, 0.979] |

